# The Obesity–MASLD–Myopenia Cascade: A Dynamic Loop within Metabolic Metamorphosis in Liver Disease

**DOI:** 10.1101/2025.09.18.25335798

**Authors:** Atsushi Nakamura, Takeshi Ichikawa, Keiji Okuyama

## Abstract

**Background & Aims:** The metabolic interplay between obesity and myopenia (MP) in chronic liver disease (CLD) remains poorly understood. We investigated whether progressive liver dysfunction mediates an obesity–MASLD (metabolic dysfunction□associated steatotic liver disease)–MP cascade and assessed the prognostic impact of MP and myopenic obesity (MO) in advanced CLD (ACLD).

**Methods:** We analyzed 859 CLD patients cross□sectionally and 169 obese patients longitudinally (median 38□months), using multimodal MRI to measure liver stiffness (LS), proton□density fat fraction (PDFF), and body composition. Temporal relationships between changes (Δ) in adiposity, muscle mass, and liver injury markers were assessed. Prognosis in ACLD (n=328) was evaluated using Cox regression.

**Results:** MP and MO were present in 29% and 8% of patients, respectively. In the longitudinal cohort, MO prevalence increased significantly from 15% to 23% (P□<□0.01). In fibrosis stages F0–2, Δvisceral adipose tissue significantly correlated with ΔPDFF, ΔALT, and ΔLS (all P□<□0.01), whereas Δmuscle mass decreased, likely from weight loss. In F3–4, ΔALBI score and ΔPDFF (hepatic fat "burning□out") independently correlated with Δmuscle mass (both P□<□0.01). In ACLD, MP—but not obesity itself—was an independent predictor of liver□related death (HR□2.27, 95%□CI□1.08–4.78, P□=□0.025).

**Conclusions:** Our findings suggest an obesity–MASLD–MP cascade driven by a liver-centered metabolic paradox: preserved hepatic function promotes harmful fat accumulation, whereas hepatic dysfunction leads to fat depletion (energy deficiency) and muscle loss. Recognition of this dynamic loop highlights the need for stage-specific strategies: fat reduction initially, followed by aggressive muscle preservation and energy repletion in ACLD.

## Introduction

Myopenic obesity (MO)―the paradoxical coexistence of muscle wasting (myopenia; MP) and obesity―is linked to poor prognosis in chronic liver disease (CLD), particularly metabolic dysfunction–associated steatotic liver disease (MASLD) [1,2]. Although MP alone predicts worse outcomes [3], whether concurrent obesity adds further risk remains unclear. MO prevalence in MASLD varies widely (5–30%) [4–6], suggesting a distinct metabolic dysfunction not captured by conventional severity scores such as the Model for End-Stage Liver Disease–sodium (MELD-Na) or albumin–bilirubin (ALBI) scores. This paradox reflects the liver’s central role in systemic energy imbalance. Recent studies [7–9] describe advanced MASLD’s “burn-out/burning-out” state—hepatic fat depletion with profound metabolic dysfunction—accelerating muscle loss and worsening MO. We therefore developed a MASLD-specific multimodal magnetic resonance imaging (MRI) system that integrates liver pathology with whole-body metabolism, enabling simultaneous measurement of liver stiffness (LS), proton density fat fraction (PDFF), muscle, and adipose tissue. Using this system, we evaluated the prognostic impact of obesity, MP, and MO in a CLD cohort through both cross-sectional and longitudinal analyses.

## Materials and Methods

### 2.1 Study Design and Population

We performed a retrospective cohort study of CLD patients who underwent liver MRI at Nippon Koukan Hospital between 2018 and 2024. A total of 859 patients with complete clinical, biochemical, and imaging data were included in the cross-sectional analysis. Additionally, to longitudinally analyze the pathophysiology of MO, a subgroup of 169 obese CLD patients who underwent repeated MRI and magnetic resonance elastography (MRE) examinations was evaluated. Demographics (age, sex, and body mass index [BMI]) and baseline laboratory values were recorded. The median interval between blood sampling and MRI was 7 days (mean, 9 ± 10 days). Patients with advanced extrahepatic malignancies or incomplete imaging data were excluded. Hepatocellular carcinoma (HCC) was not an exclusion criterion for cross-sectional analysis but was excluded in the longitudinal study. None of the patients were receiving glucagon-like peptide-1 receptor agonists (GLP-1 RAs) at baseline. The study adhered to the Declaration of Helsinki and was approved by the institutional review board (No. 202014); the requirement for written informed consent was waived due to its retrospective design.

### 2.2 Disease Classification, Staging, and Imaging Measurements

CLD was categorized as MASLD or non-MASLD according to the international consensus statement [2]. After an overnight fast, patients underwent 1.5-T MRI (GE Healthcare), including MRE for LS and IDEAL-IQ for PDFF to quantify hepatic fat. Fibrosis (F) staging was based on etiology-specific MRE-LS cutoffs for MASLD [10] and non-MASLD [11].

**-** Cirrhosis was subclassified as compensated (F4c) or decompensated (F4d) per the 2018 EASL guidelines [12].
**-** Liver function was assessed by ALBI and MELD-Na scores [13,14].
**-** Prognosis was analyzed only in patients with ACLD (LS ≥ 3.0 kPa), in line with the AASLD definition [15] and supported by our previous reports [16].

Because no universally accepted definition of “burn-out” in MASLD exists, we defined “burning-out” operationally as marked hepatic fat reduction (PDFF < 5% [7]) in patients with ACLD who had previously documented hepatic steatosis by biopsy or imaging. Such hepatic fat reduction in ACLD patients with MASLD has been reported to be associated with a poor prognosis [7–9].

### 2.3 Definition of Obesity and MP

Obesity was defined according to WHO/IASO/IOTF Asian criteria as BMI ≥ 25 kg/m² [17]. MRI was chosen to simultaneously quantify skeletal muscle mass, hepatic fat, and LS (**Supplementary Methods, Supplementary Figure 1**).

MP was diagnosed using the MRI-derived paraspinal muscle index (PSMI) at the level of the superior mesenteric artery (SMA) [18–20]. MO was defined as concurrent obesity and MP. Visceral (VAT) and subcutaneous (SAT) adipose tissue areas were measured on a single axial T2-weighted slice at the SMA level. Steatosis is strongly influenced by mesenteric VAT (mVAT), which is directly connected to the portal circulation; therefore, both VAT and SAT were assessed at the SMA level [21,22]. This standardized slice-matching protocol provided robust, reproducible annualized percent changes (Δ%/year) with minimal positional variability in longitudinal analysis (see **Supplementary Figure 1**). VAT could not be assessed in patients with overt ascites; analyses were therefore limited to compensated cases.

### 2.4 Outcome and Follow-up

Patients with ACLD were followed until liver-related death, liver transplantation, or last confirmed contact. Liver-related mortality included deaths from hepatic decompensation, variceal bleeding, spontaneous bacterial peritonitis, or HCC. Follow-up data were collected through medical records, interinstitutional communications, or direct contact.

Follow-up for the ACLD prognostic cohort (n=328) continued until liver-related death, liver transplantation, or last contact; the median clinical follow-up was 34 months (interquartile range: 3–73 months). For the longitudinal obese-CLD cohort without HCC (n=169), the median imaging follow-up between serial MRE assessments was 38 months.

*Further details regarding imaging protocols, fibrosis staging criteria, muscle mass assessment, and follow-up procedures are provided in the **Supplementary Materials***.

### 2.5 Statistical Analysis

Statistical analyses were performed using JMP version 18.2.0 (SAS Institute Japan). Between-group differences were assessed using parametric (Student’s t-test) or non-parametric (Mann–Whitney U test, Wilcoxon signed-rank test) methods, as appropriate. Variables showing p < 0.05 in univariate analysis were considered candidates for multivariate analyses. Stepwise forward and backward selection methods based on Akaike Information Criterion (AIC) were used to refine candidate variables. Subsequently, a minimal set of clinically relevant variables identified by this process was included in the final multivariate logistic regression and Cox proportional hazards models. Survival was assessed using Kaplan–Meier curves and log-rank tests. A p-value < 0.05 was considered statistically significant.

For the primary prognostic analysis, no adjustment for multiple comparisons was applied due to its hypothesis-driven nature. However, the exploratory correlation analyses involving skeletal muscle mass, VAT, PDFF, and ALBI score over the follow-up period were adjusted using the False Discovery Rate (FDR) method (Benjamini–Hochberg procedure) to control for multiple testing. FDR-adjusted P-values < 0.05 were considered statistically significant for these analyses.

*Detailed methods are provided in the **Supplementary Material**s*.

### 2.6 Use of AI-based tools

Portions of this manuscript were generated with the assistance of ChatGPT (OpenAI) and were thoroughly reviewed and edited by the authors.

## Results

### Clinical characteristics (Table 1)

Among 859 patients, 245 (29%) had MP, including 64 (8%) with MO; the remaining 614 were non-MP. MP patients were older and predominantly male. MASLD and alcohol-related liver disease were common in MO. Compared with non-MP, MP patients had worse nutritional status, higher inflammation, and greater disease severity; MP-alone and MO showed similar severity. Muscle mass was equally reduced in both MP groups, yet BMI and hepatic fat (PDFF) were lowest in MP-alone. Diabetes prevalence did not differ among groups.

**Table 1.**
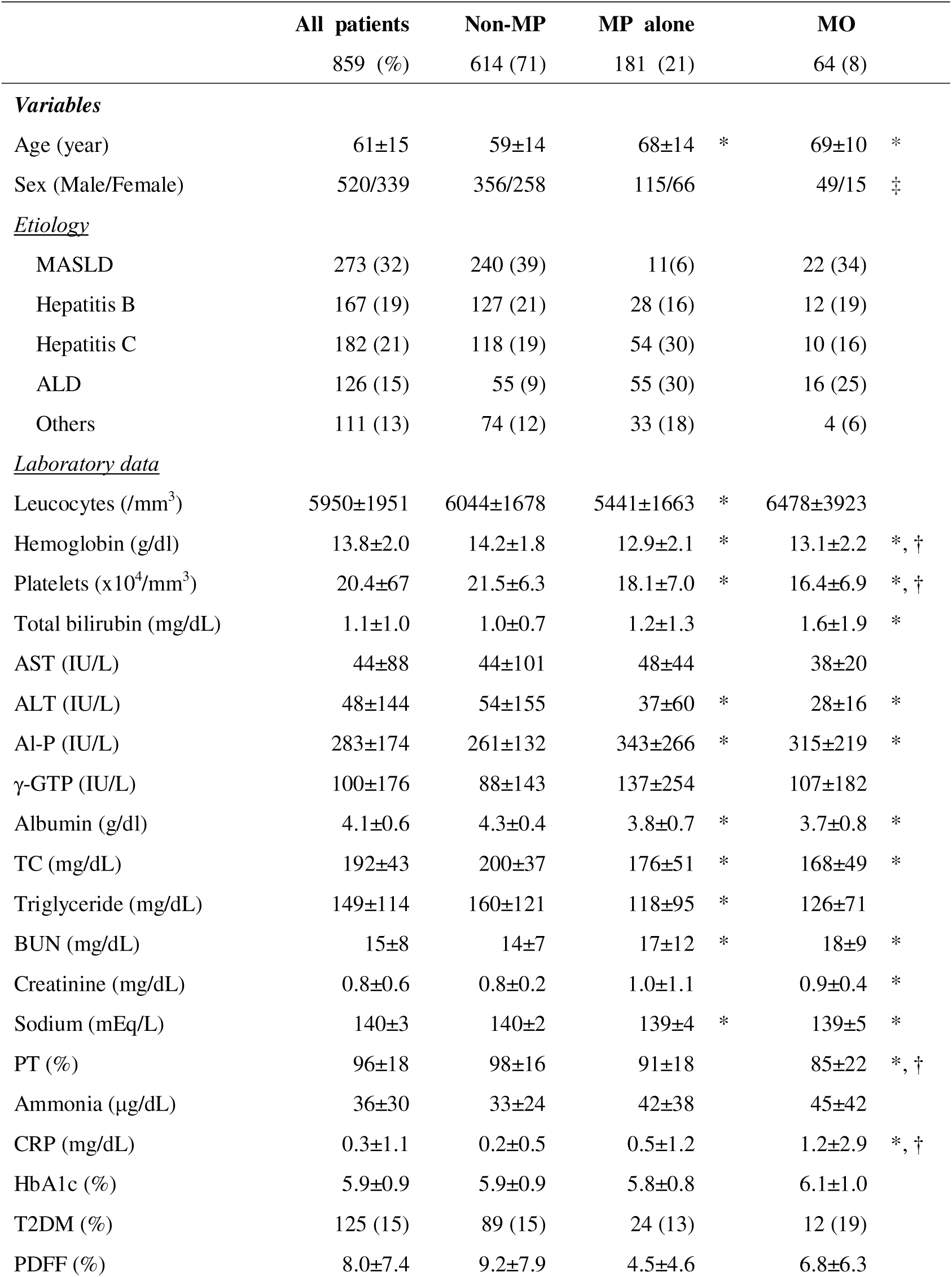
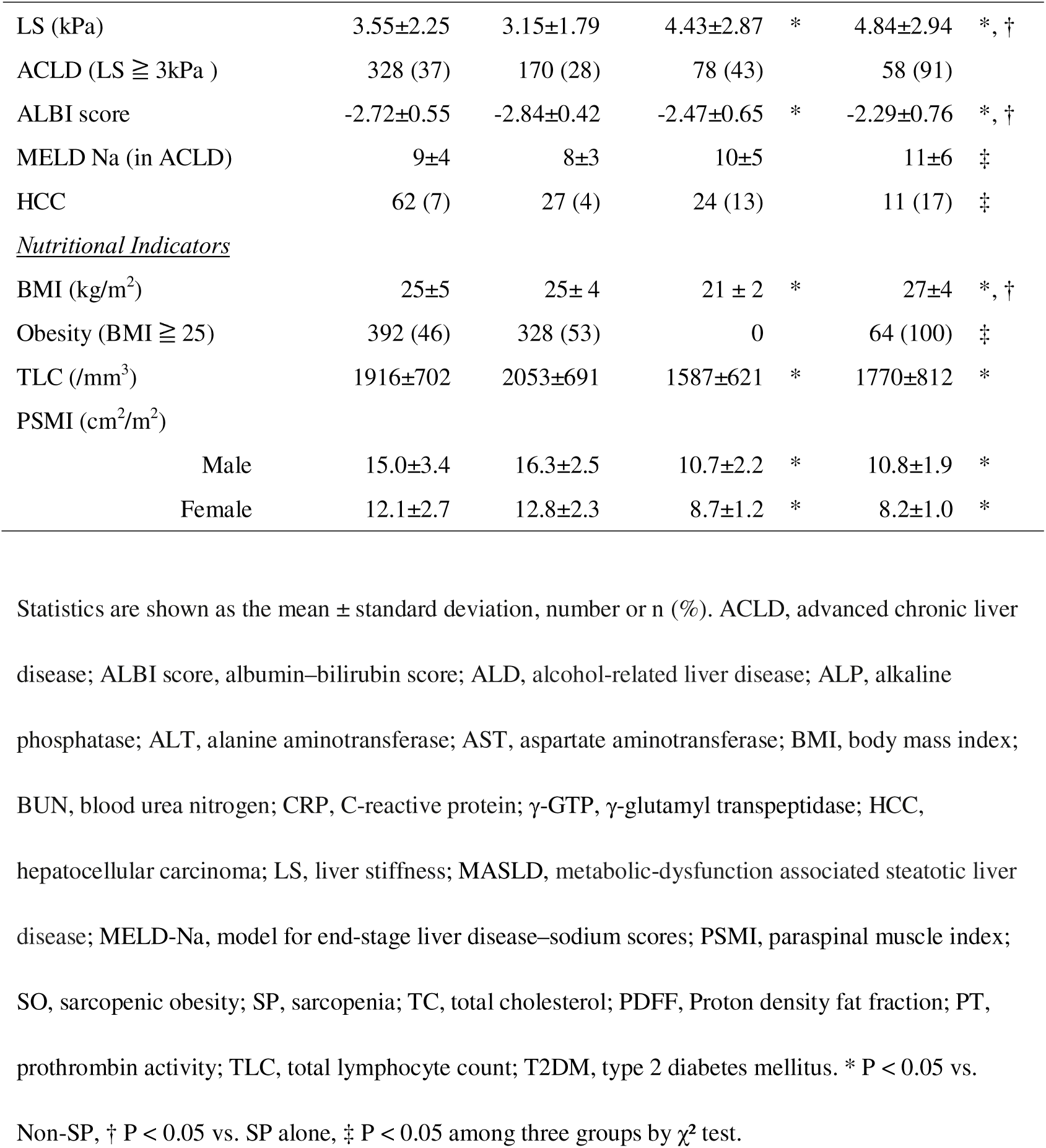
Clinical characteristics of the study population stratified by Myopenia (MP) and obesity status.

|  | All patients<br>859 (%) | Non-MP<br>614 (71) | MP alone<br>181 (21) | MO<br>64 (8) |
| --- | --- | --- | --- | --- |
| ***Variables*** |  |  |  |  |
| Age (year) | 61±15 | 59±14 | 68±14 * | 69±10 * |
| Sex (Male/Female) | 520/339 | 356/258 | 115/66 | 49/15 ‡ |
| ***Etiology*** |  |  |  |  |
| MASLD | 273 (32) | 240 (39) | 11(6) | 22 (34) |
| Hepatitis B | 167 (19) | 127 (21) | 28 (16) | 12 (19) |
| Hepatitis C | 182 (21) | 118 (19) | 54 (30) | 10 (16) |
| ALD | 126 (15) | 55 (9) | 55 (30) | 16 (25) |
| Others | 111 (13) | 74 (12) | 33 (18) | 4 (6) |
| ***Laboratory data*** |  |  |  |  |
| Leucocytes (/mm<sup>3</sup>) | 5950±1951 | 6044±1678 | 5441±1663 * | 6478±3923 |
| Hemoglobin (g/dl) | 13.8±2.0 | 14.2±1.8 | 12.9±2.1 * | 13.1±2.2 *, † |
| Platelets (x10<sup>4</sup>/mm<sup>3</sup>) | 20.4±67 | 21.5±6.3 | 18.1±7.0 * | 16.4±6.9 *, † |
| Total bilirubin (mg/dL) | 1.1±1.0 | 1.0±0.7 | 1.2±1.3 | 1.6±1.9 * |
| AST (IU/L) | 44±88 | 44±101 | 48±44 | 38±20 |
| ALT (IU/L) | 48±144 | 54±155 | 37±60 * | 28±16 * |
| Al-P (IU/L) | 283±174 | 261±132 | 343±266 * | 315±219 * |
| γ-GTP (IU/L) | 100±176 | 88±143 | 137±254 | 107±182 |
| Albumin (g/dl) | 4.1±0.6 | 4.3±0.4 | 3.8±0.7 * | 3.7±0.8 * |
| TC (mg/dL) | 192±43 | 200±37 | 176±51 * | 168±49 * |
| Triglyceride (mg/dL) | 149±114 | 160±121 | 118±95 * | 126±71 |
| BUN (mg/dL) | 15±8 | 14±7 | 17±12 * | 18±9 * |
| Creatinine (mg/dL) | 0.8±0.6 | 0.8±0.2 | 1.0±1.1 | 0.9±0.4 * |
| Sodium (mEq/L) | 140±3 | 140±2 | 139±4 * | 139±5 * |
| PT (%) | 96±18 | 98±16 | 91±18 | 85±22 *, † |
| Ammonia (μg/dL) | 36±30 | 33±24 | 42±38 | 45±42 |
| CRP (mg/dL) | 0.3±1.1 | 0.2±0.5 | 0.5±1.2 | 1.2±2.9 *, † |
| HbA1c (%) | 5.9±0.9 | 5.9±0.9 | 5.8±0.8 | 6.1±1.0 |
| T2DM (%) | 125 (15) | 89 (15) | 24 (13) | 12 (19) |
| PDFF (%) | 8.0±7.4 | 9.2±7.9 | 4.5±4.6 | 6.8±6.3 |

|  |  |  |  |  |  |  |
| --- | --- | --- | --- | --- | --- | --- |
| LS (kPa) | 3.55±2.25 | 3.15±1.79 | 4.43±2.87 | * | 4.84±2.94 | *, † |
| ACLD (LS ≥ 3kPa ) | 328 (37) | 170 (28) | 78 (43) |  | 58 (91) |  |
| ALBI score | -2.72±0.55 | -2.84±0.42 | -2.47±0.65 | * | -2.29±0.76 | *, † |
| MELD Na (in ACLD) | 9±4 | 8±3 | 10±5 |  | 11±6 | ‡ |
| HCC | 62 (7) | 27 (4) | 24 (13) |  | 11 (17) | ‡ |
| *Nutritional Indicators* |  |  |  |  |  |  |
| BMI (kg/m<sup>2</sup>) | 25±5 | 25±4 | 21±2 | * | 27±4 | *, † |
| Obesity (BMI ≥ 25) | 392 (46) | 328 (53) | 0 |  | 64 (100) | ‡ |
| TLC (/mm<sup>3</sup>) | 1916±702 | 2053±691 | 1587±621 | * | 1770±812 | * |
| PSMI (cm<sup>2</sup>/m<sup>2</sup>) |  |  |  |  |  |  |
| Male | 15.0±3.4 | 16.3±2.5 | 10.7±2.2 | * | 10.8±1.9 | * |
| Female | 12.1±2.7 | 12.8±2.3 | 8.7±1.2 | * | 8.2±1.0 | * |

Statistics are shown as the mean ± standard deviation, number or n (%). ACLD, advanced chronic liver disease; ALBI score, albumin–bilirubin score; ALD, alcohol-related liver disease; ALP, alkaline phosphatase; ALT, alanine aminotransferase; AST, aspartate aminotransferase; BMI, body mass index; BUN, blood urea nitrogen; CRP, C-reactive protein; $\gamma$ -GTP, $\gamma$ -glutamyl transpeptidase; HCC, hepatocellular carcinoma; LS, liver stiffness; MASLD, metabolic-dysfunction associated steatotic liver disease; MELD-Na, model for end-stage liver disease–sodium scores; PSMI, paraspinal muscle index; SO, sarcopenic obesity; SP, sarcopenia; TC, total cholesterol; PDFF, Proton density fat fraction; PT, prothrombin activity; TLC, total lymphocyte count; T2DM, type 2 diabetes mellitus. \* P < 0.05 vs. Non-SP, † P < 0.05 vs. SP alone, ‡ P < 0.05 among three groups by $\chi^2$ test.

### Prevalence of Obesity, MP and MO

Etiology□specific prevalence — Obesity was 78% in MASLD vs 31% in non-MASLD, whereas MP was 12% vs 32% (both P < 0.001); MO rates were similar (8% vs 7%, P = 0.566).

**Figure 1** shows etiology-specific trends in obesity, MP, and MO across fibrosis stages (F0–2, F2–3, F4c, F4d).

- MASLD (panels A–C): Prevalence of all phenotypes rises step-wise, peaking at decompensated cirrhosis.
- Non-MASLD (panels D–F): Obesity peaks at compensated cirrhosis then declines, MP steadily increases, and MO remains consistently low.

**Figure 1.**
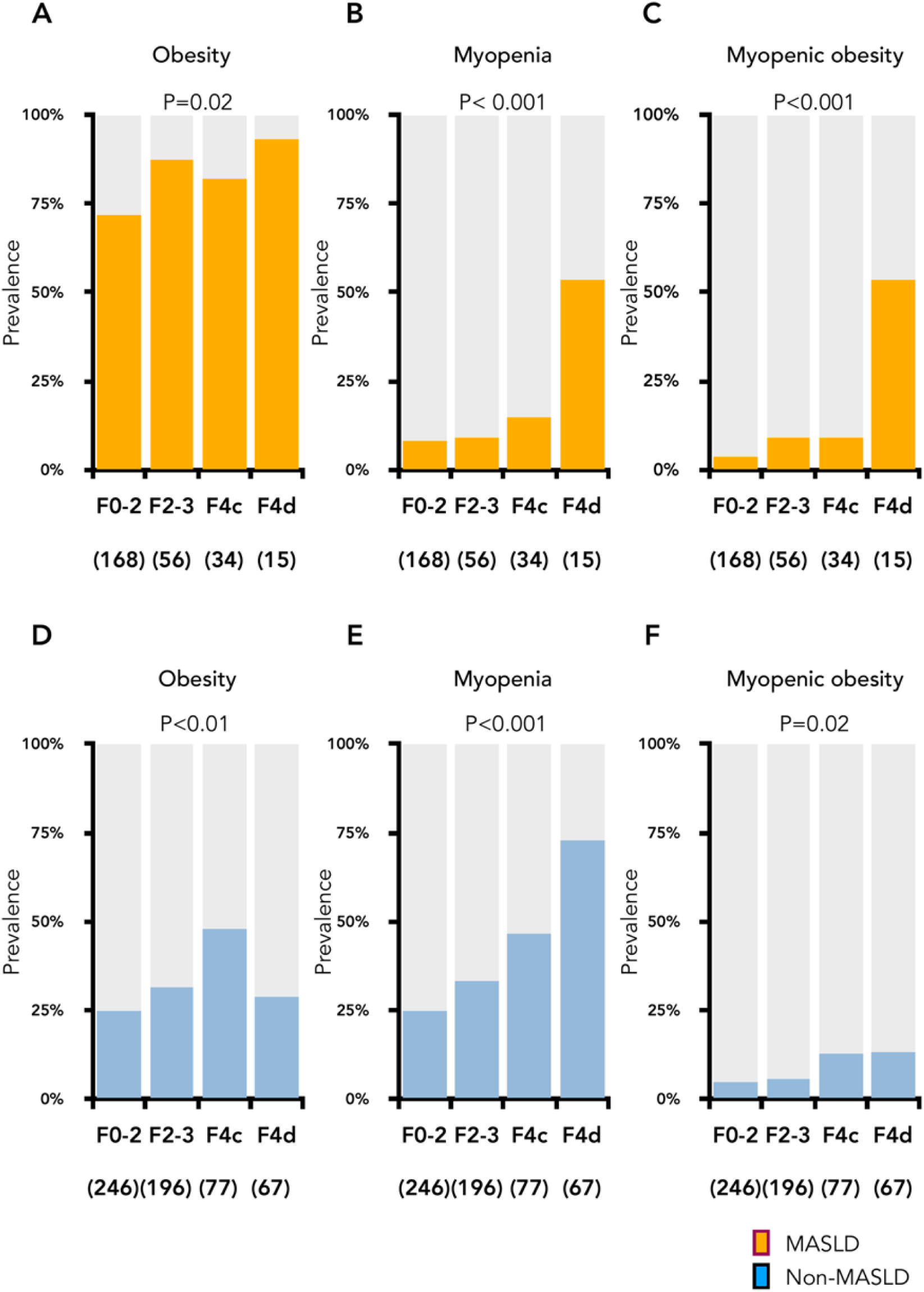
Prevalence of obesity, myopenia (MP), and myopenic obesity (MO) by fibrosis stage in MASLD (A–C) and non-MASLD (D–F). MASLD = orange, non-MASLD = blue.

These divergent trajectories highlight distinct disease progression patterns in MASLD and non-MASLD.

**Figure 2** depicts trajectories of PDFF, alanine aminotransferase (ALT), and ALBI across fibrosis stages (values in **Supplementary Table 1**).

- MASLD (panels A–C): PDFF and ALT rise from early to intermediate fibrosis but drop sharply at F4d, illustrating the "burn-out" phenotype. ALBI worsens progressively, with a marked increase at F4d.
- Non-MASLD (panels D–F): PDFF remains low across all stages, ALT steadily increases before slightly declining at F4d, and ALBI deteriorates progressively in advanced fibrosis.

**Figure 2.**
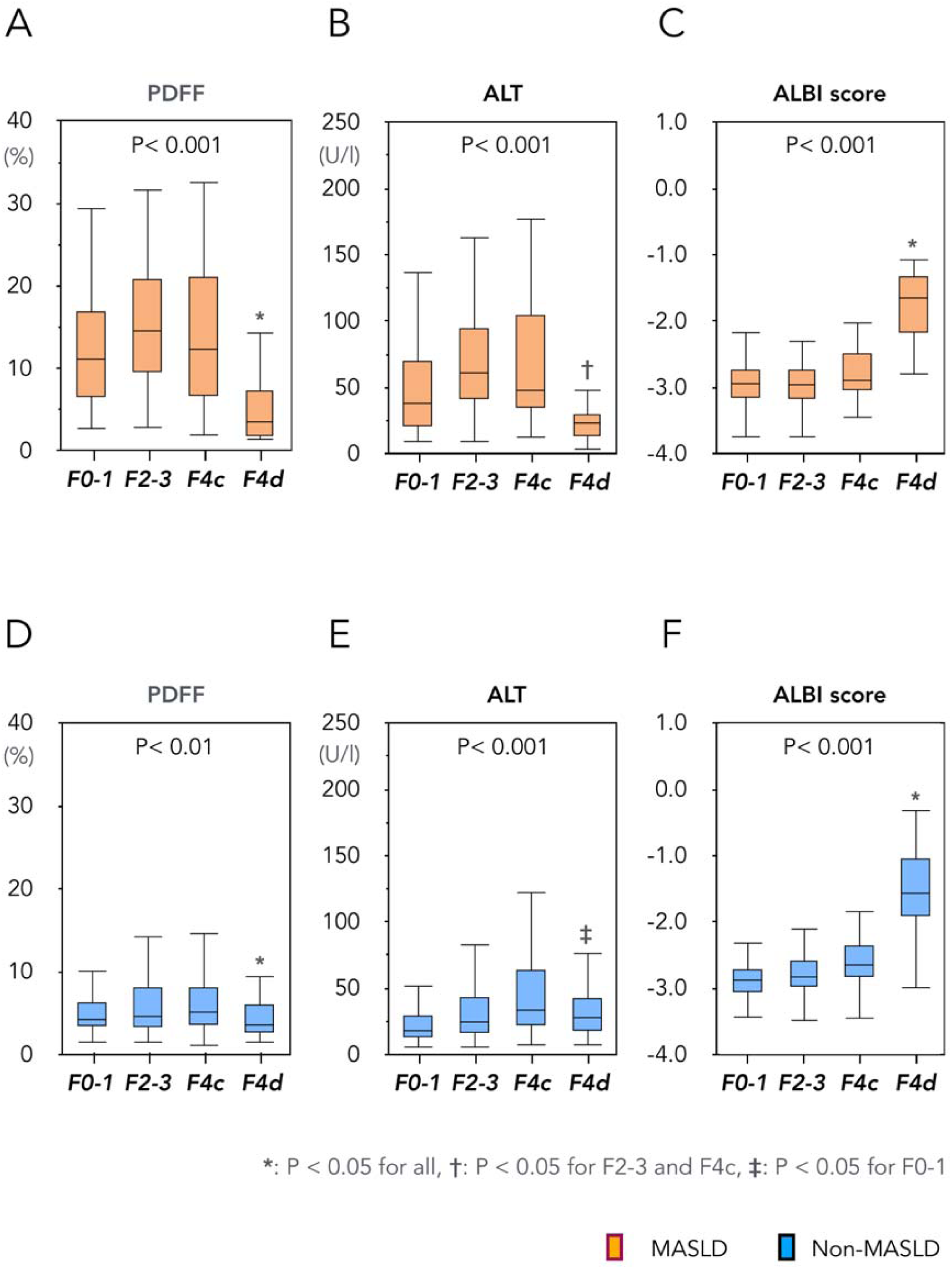
Changes in PDFF (A, D), ALT (B, E), and ALBI score (C, F) by fibrosis stage in MASLD and non-MASLD groups. MASLD = orange, non-MASLD = blue.

These differences underscore distinct metabolic and functional shifts during fibrosis progression in MASLD versus non-MASLD.

**Supplementary Figure 2** shows that in F3–4 (without HCC), ALBI score correlated positively with LS in both groups. However, the negative correlation between ALBI and PDFF differed depending on etiology: non-MASLD maintained low PDFF, whereas MASLD showed marked PDFF decline with decreasing liver functional reserve. Both groups ultimately converged toward a common metabolic endpoint of hepatic fat loss.

### Longitudinal Analysis of Obese Patients in CLD without HCC

Longitudinal analysis included 169 obese CLD patients without HCC who underwent repeat MRE assessments (median follow-up 38 months; **Supplementary Table 2**).

During follow-up, all medications were administered for ≥ 3 months: 30 patients received antidiabetic agents (GLP-1 RAs = 7, SGLT2 inhibitors = 28, with overlap). All MASLD patients were advised to lose weight; 52% also received dietitian-led counselling (structured counselling in 37% of the total cohort). At baseline, MASLD patients were younger and had higher BMI, ALT, and PDFF compared to non-MASLD patients (all P < 0.05).

During follow-up, modest improvements were observed in ALT, PDFF, and LS; however, muscle mass declined significantly (**Figure 3**). The prevalence of MP also increased significantly from 15% to 23% (**Figure 4A**; P = 0.009).

**Figure 3.**
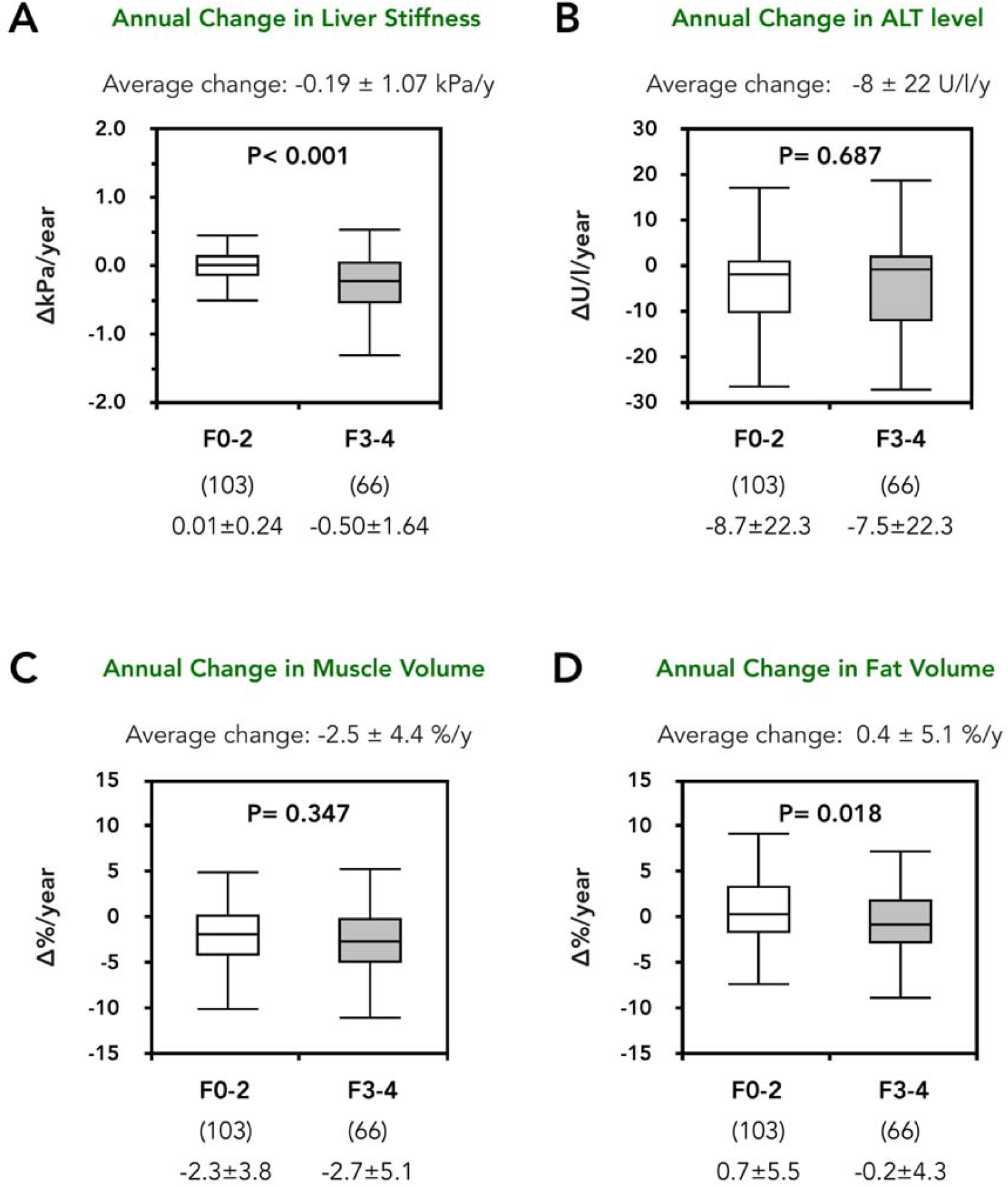
Longitudinal changes in liver stiffness (A), ALT (B), muscle volume (C), and fat volume (D) in obese CLD patients without HCC. Values are mean ± SD.

**Figure 4.**
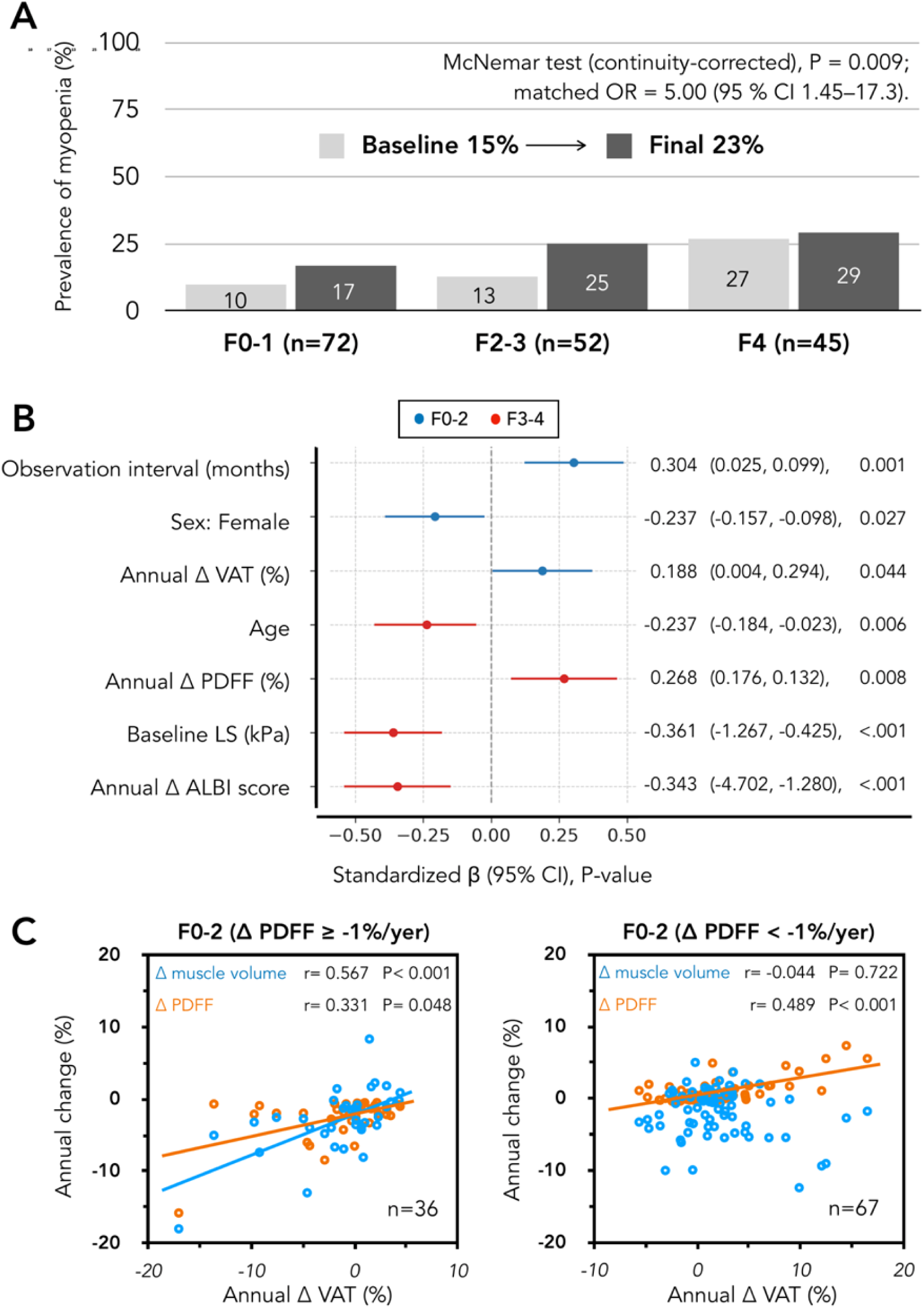
Longitudinal prevalence and risk factors of myopenia (MP) among obese patients with chronic liver disease (CLD) without hepatocellular carcinoma (HCC). **(A)** Prevalence of MP at baseline and follow-up in obese CLD patients. *P=0.009 (McNemar test). **(B)** Multivariate analysis of factors for annual muscle mass change by fibrosis stage (F0–2, blue; F3–4, red). Values are standardized β with 95% CIs. **(C)** Correlation between ΔVAT and muscle/PDFF in F0–2. In patients with hepatic fat reduction (ΔPDFF ≥ 1%/yr), VAT decrease was associated with PDFF reduction and paradoxical muscle loss. In those with stable or increased PDFF (ΔPDFF < 1%/yr),

#### I. Analysis of risk factors for MP in obese CLD

To clarify factors underlying MP, we analyzed correlations between annual muscle-volume changes and parameters of liver pathology, hepatic fat, and body composition, stratified by fibrosis stages (F0–2: n = 103, F3–4: n = 66).

In the overall F0–2 group, annual muscle-volume changes were significantly associated only with annual changes in mesenteric VAT (mVAT), not with total adipose tissue (**Supplementary Table 3**), a finding that held true in the MASLD subset (**Supplementary Table 4**, n=63). To investigate the broader role of mVAT as a metabolic driver, we then conducted a comprehensive response screening analysis. The analysis revealed that annual mVAT change was most strongly correlated with the annual change in PDFF (R²=0.40, FDR-adjusted P < 0.001). Furthermore, ALT (FDR-adjusted P < 0.001) and muscle mass (FDR-adjusted P = 0.048) changes also showed significant correlations after FDR correction. These findings identify mVAT as a central metabolic hub in early-stage MASLD, simultaneously linked to the progression of hepatic steatosis, and inflammation.

In F3–4 (**Supplementary Table 5**), worsening ALBI scores were moderately and significantly correlated with declines in both skeletal muscle volume and body fat mass. ALBI worsening was also significantly correlated with reduced annual ΔPDFF, supporting a "burning-out" process. These findings were consistent with our longitudinal analysis.

We then conducted multivariate analysis stratified by fibrosis stage (F0–2 and F3–4), including age, sex, BMI, liver function tests, PDFF, LS, ALBI score, body composition, concomitant medications, receipt of nutritional counseling, and decompensated cirrhosis as explanatory variables. Significant factors identified by univariate analysis (**Supplementary Table 6**) were as follows: in F0–2, sex, BUN, Cr, annual ΔVAT, and observation interval; in F3–4, age, annual ΔPDFF, baseline LS, baseline ALBI score, and annual ΔALBI score.

**Figure 4B** summarizes the multivariate analysis. In F0–2, annual ΔVAT emerged as an independent predictor of annual muscle-volume change, alongside follow-up interval and female sex, confirming a direct muscle-fat coupling. In stark contrast, for F3–4, the significant predictors shifted entirely to markers of liver dysfunction and aging: age, annual ΔPDFF, baseline LS, and annual ΔALBI score. Notably, annual ALBI worsening strongly correlated with muscle loss (FDR-adjusted P < 0.001), and annual hepatic fat reduction (ΔPDFF; "burning-out") similarly associated with muscle decline (FDR-adjusted P < 0.001).

The independent effect of ΔVAT likely reflects the strong muscle–fat coupling observed in the MASLD subgroup achieving ≥ 1%/year PDFF reduction (**Figure 4C**), even though associations were weaker in the overall F0–2 cohort. Collectively, these findings highlight mVAT as a modifiable driver of disease progression and paradoxical muscle loss in obese CLD, particularly MASLD.

#### II. Subtypes of MO in CLD

Three fibrosis-stage–specific metabolic phenotypes emerged (Figure 4–5):

1. Starvation-like type (F0–2), driven by dietary restriction (**Figure 4C**).
2. Burning-out type (advanced MASLD), characterized by marked hepatic fat loss (**Figure 5A–D**).
3. Liver-failure type (cirrhosis), associated with impaired hepatic function (**Figure 5E–F**).

**Figure 5.**
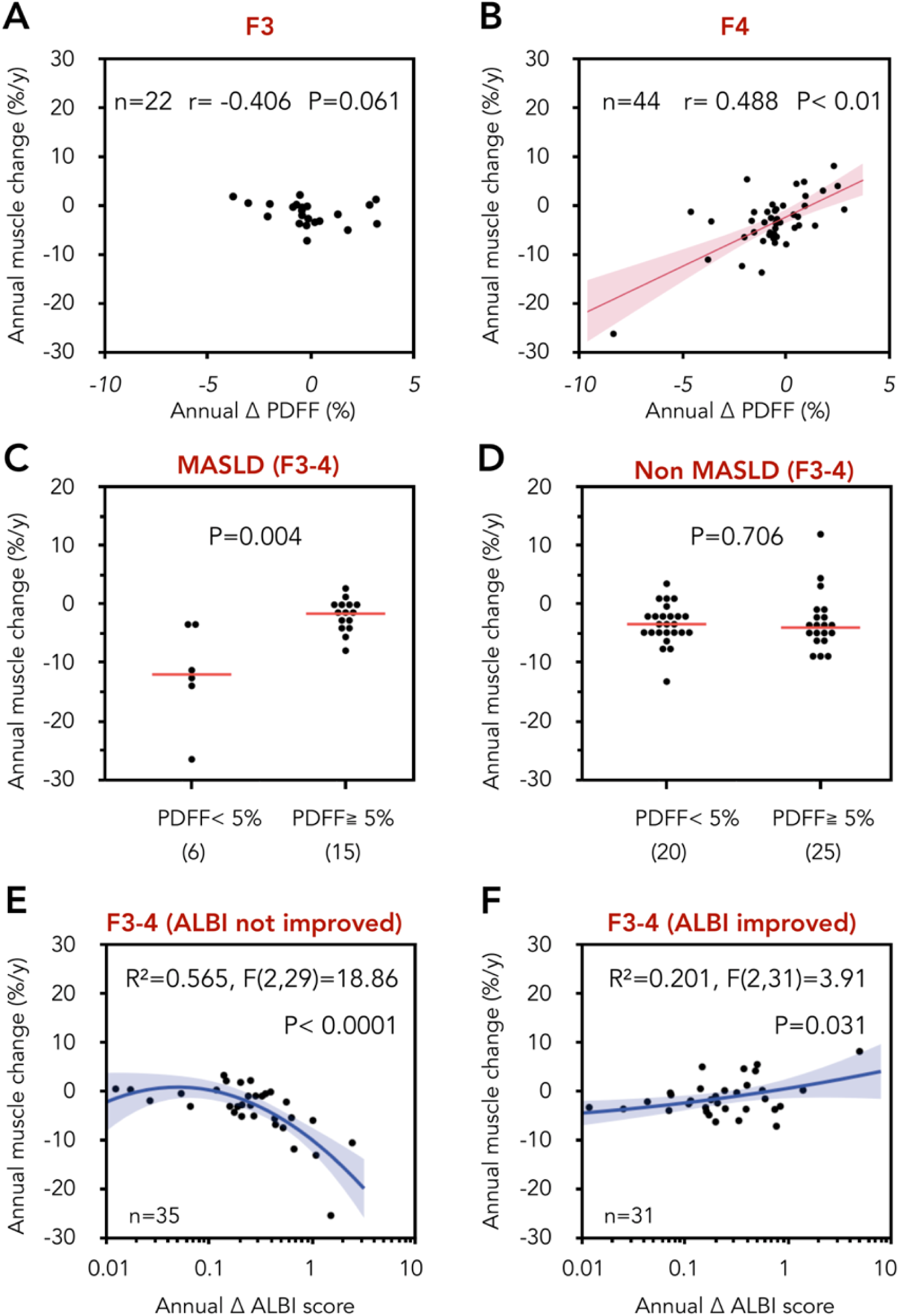
Relationship between changes in hepatic fat (PDFF), ALBI scores, and annual muscle change in patients with advanced fibrosis (F3-4). **(A, B)** Correlation between ΔPDFF and muscle mass in F3 (A) and F4 (B). No significant association was seen in F3, where portal hypertension is the main driver of progression. In F4, a significant positive association was observed. **(C, D)** Annual muscle mass change by liver fat status (PDFF < 5% vs ≥ 5%) in MASLD and non-MASLD (F3–4). In MASLD, low fat (PDFF < 5%)—the “burning-out” phenotype, reflecting severely impaired hepatic reserve—was associated with accelerated muscle loss, with no difference in non-MASLD. **(E, F)** Correlation between annual changes in hepatic functional reserve (absolute ΔALBI score) and muscle mass in patients with no improvement (E, ΔALBI ≥ –0.01) or improvement (F, ΔALBI < –0.01). Muscle loss was pronounced with no improvement, whereas improvement was associated with only modest gain. Regression lines with 95% CIs are shown.

### Prognostic impact of MP and MO in ACLD

Among 328 ACLD patients (mean follow-up, 34 ± 18 months; median, 34.0 months), 56 liver-related events occurred: 38 liver failures, 18 HCCs, and two liver transplants. Patients were categorized into four groups by MP and obesity status; survival was compared via Kaplan–Meier analysis (**Figure 6A**). MP was associated with significantly poorer survival, irrespective of obesity (log-rank P < 0.001), while prognosis did not differ between MP-only and MO.

**Figure 6.**
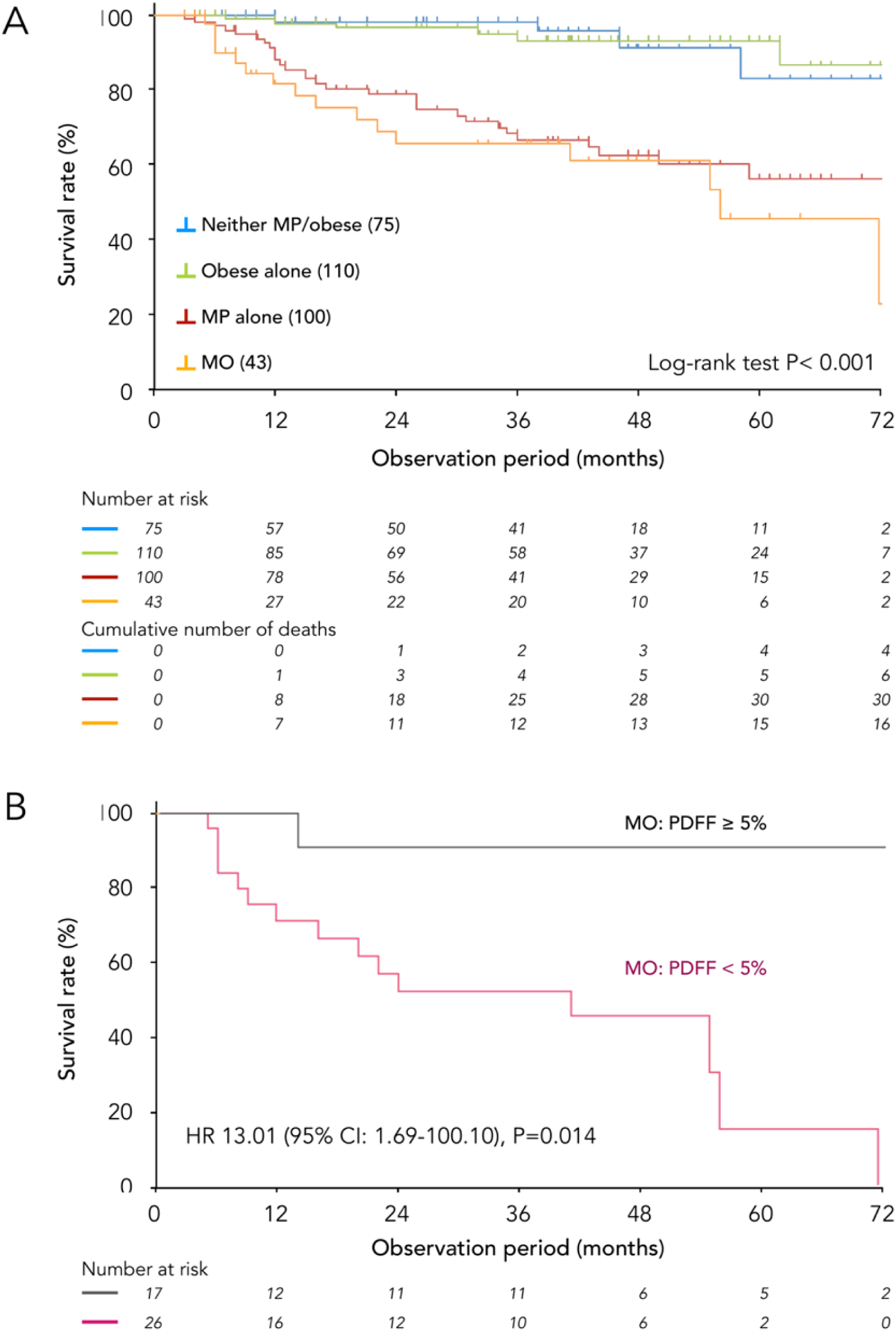
**(A)** Kaplan–Meier survival analysis in ACLD by myopenia (MP) and obesity status. Log-rank P < 0.001. **(B)** Subgroup survival analysis in myopenic obesity (MO) by PDFF (< 5% vs. ≥ 5%).

In univariate analysis (**Table 2**), 18 factors were prognostic. Of these, 4 were selected for multivariate analysis using stepwise selection (both directions) and clinical relevance. Multivariate analysis identified ALBI score, HCC, and MP as independent prognostic factors. The model showed excellent fit (likelihood-ratio χ² = 109.4, df = 5, P < 0.001), high events-per-variable ratio: 19, and satisfied proportional hazards (global Schoenfeld P = 0.65).

**Table 2.**
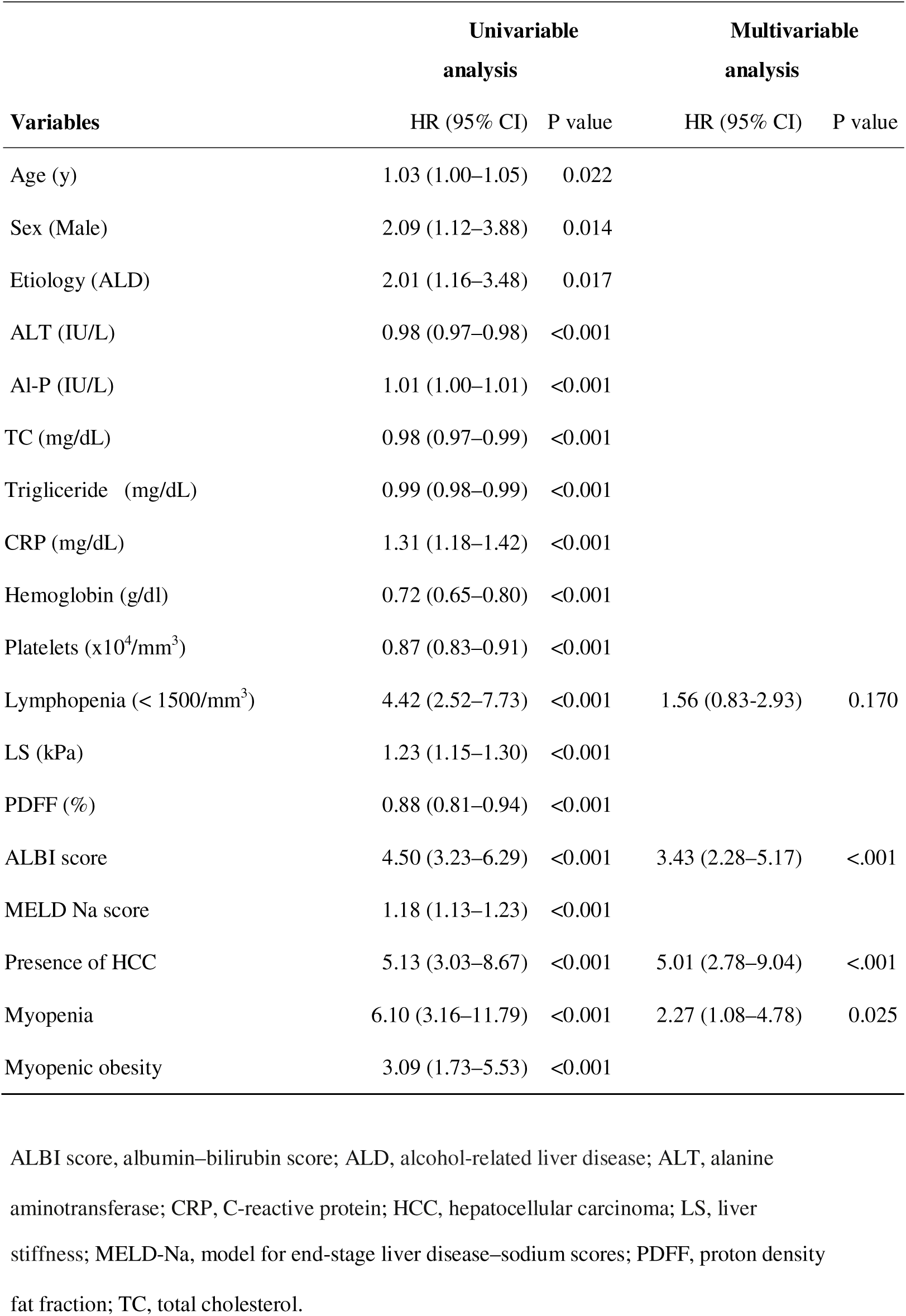
Univariate and multivariate Cox proportional hazards analysis of factors associated with liver-related mortality in patients with advanced chronic liver disease (ACLD).

| Variables | Univariable analysis |  | Multivariable analysis |  |
| --- | --- | --- | --- | --- |
|  | HR (95% CI) | P value | HR (95% CI) | P value |
| Age (y) | 1.03 (1.00–1.05) | 0.022 |  |  |
| Sex (Male) | 2.09 (1.12–3.88) | 0.014 |  |  |
| Etiology (ALD) | 2.01 (1.16–3.48) | 0.017 |  |  |
| ALT (IU/L) | 0.98 (0.97–0.98) | <0.001 |  |  |
| Al-P (IU/L) | 1.01 (1.00–1.01) | <0.001 |  |  |
| TC (mg/dL) | 0.98 (0.97–0.99) | <0.001 |  |  |
| Triglyceride (mg/dL) | 0.99 (0.98–0.99) | <0.001 |  |  |
| CRP (mg/dL) | 1.31 (1.18–1.42) | <0.001 |  |  |
| Hemoglobin (g/dl) | 0.72 (0.65–0.80) | <0.001 |  |  |
| Platelets ($\times 10^4/\text{mm}^3$) | 0.87 (0.83–0.91) | <0.001 | | |
| Lymphopenia ($< 1500/\text{mm}^3$) | 4.42 (2.52–7.73) | <0.001 | 1.56 (0.83–2.93) | 0.170 |
| LS (kPa) | 1.23 (1.15–1.30) | <0.001 |  |  |
| PDFF (%) | 0.88 (0.81–0.94) | <0.001 |  |  |
| ALBI score | 4.50 (3.23–6.29) | <0.001 | 3.43 (2.28–5.17) | <.001 |
| MELD Na score | 1.18 (1.13–1.23) | <0.001 |  |  |
| Presence of HCC | 5.13 (3.03–8.67) | <0.001 | 5.01 (2.78–9.04) | <.001 |
| Myopenia | 6.10 (3.16–11.79) | <0.001 | 2.27 (1.08–4.78) | 0.025 |
| Myopenic obesity | 3.09 (1.73–5.53) | <0.001 |  |  |

ALBI score, albumin–bilirubin score; ALD, alcohol-related liver disease; ALT, alanine aminotransferase; CRP, C-reactive protein; HCC, hepatocellular carcinoma; LS, liver
stiffness; MELD-Na, model for end-stage liver disease–sodium scores; PDFF, proton density fat fraction; TC, total cholesterol.

### Subgroup analysis of MO: impact of hepatic fat content

A subgroup analysis was performed on patients with MO (**Figure 6B**), stratified by hepatic fat content. Patients with sufficient hepatic fat (PDFF ≥ 5%, n = 17) showed excellent survival, whereas those with depleted hepatic fat (burn-out type, PDFF < 5%, n = 26) had significantly poorer outcomes. These findings suggest that hepatic fat, once a driver of early MASLD, may later act as an energy source in malnourished ACLD.

## Discussion

This multimodal MRI study proposes a novel pathophysiological model in CLD: the “obesity–MASLD–MP cascade,” defined by a dynamic metabolic shift we term the “meta-paradox.” In this framework, hepatic fat accumulation may drive disease progression in early MASLD, while in advanced cirrhosis, its depletion (“burning-out”) may reflect metabolic collapse and trigger accelerated muscle loss. This transition from energy surplus to systemic energy deficit contributes to MP and may influence clinical outcomes.

### 1. Obesity–MASLD–myopenia cascade

Our longitudinal study over three years demonstrated that MP is a progressive and largely irreversible condition once established, underscoring the clinical importance of early intervention. Traditional pathogenic factors—such as insulin resistance, fibrosis, portal hypertension, hormonal imbalance, and systemic inflammation—have typically been considered separately, lacking an integrated perspective [3,4,5]. Here, we propose an integrated “obesity–MASLD–myopenia cascade” in which obesity initiates liver dysfunction and fibrosis progression, subsequently impairing hepatic energy metabolism and muscle protein synthesis while accelerating muscle degradation—ultimately resulting in MP. Of note, this cascade represents a disease-driven phenomenon fundamentally distinct from age-related MO. While age-related MO typically involves relatively preserved metabolic crosstalk among adipose tissue, liver, and skeletal muscle, the obesity–MASLD–myopenia cascade is uniquely characterized by progressive and irreversible hepatic metabolic collapse as the primary driver of systemic muscle catabolism.

Supporting this concept, Koo et al. [23] reported that in a biopsy-proven MASLD cohort, MP was significantly more prevalent in patients with steatohepatitis (35%) than in those without (18%), and was independently associated with advanced fibrosis, regardless of BMI and insulin resistance. These findings validate the cascade, providing new insights into MASLD pathogenesis and therapeutic targets.

### 2. Metabolic Metamorphosis: From Burn-up to Burning-out

The main finding of this study is a dynamic transformation in liver metabolism from early-stage hepatic fat accumulation and inflammation (“burn-up”), driven by body fat—particularly mesenteric VAT (mVAT)—to hepatic fat depletion (“burning-out”) in advanced fibrosis and cirrhosis.

In our cross-sectional analysis, reduced PDFF in late-stage MASLD correlated with decreased ALT (indicating diminished hepatocellular inflammation and loss of viable hepatocytes [24]) and worsening ALBI scores. This suggests that inflammatory hepatocellular damage initially drives the burn-up process, subsequently progressing to burn-out through loss of functioning hepatocytes. Given the liver’s central role in nutrient metabolism, this hepatic metabolic metamorphosis shifts systemic metabolism from an anabolic to a catabolic state, ultimately resulting in muscle mass loss.

In advanced MASLD and cirrhosis, previous studies have firmly linked muscle loss to advanced fibrosis, impaired hepatic synthetic capacity, portal hypertension, hormonal dysregulation, and systemic inflammation [2]. In our cohort, muscle mass was already reduced in the early stage (F0–2), yet this decline showed no clear association with conventional liver-dysfunction or fibrosis markers (ΔALT, ΔLS).

Consistent with earlier reports, adipose tissue—particularly mVAT—was the factor most strongly associated with early liver-disease progression [21,22,25] and glycemic control (HbA1c) in F0–2. Given the direct portal drainage of mVAT-derived metabolites to the liver, this VAT-first pattern is biologically plausible and underscores the sensitivity of our multimodal Δ-analysis in detecting early metabolic signals. Indeed, insulin resistance and chronic inflammation originating from adipose tissue have previously been identified as critical factors contributing to MASLD-associated MP [26, 27]. Therefore, our findings suggest that obesity may represent a common upstream factor potentially contributing to both liver dysfunction and muscle loss from the early stage of MASLD.

The proposed cascade highlights a clinical “meta-paradox”: hepatic fat initially accelerates disease but its later depletion signals poor prognosis [7]. Our data—even with a modest number of MO cases—suggest that preserved steatosis may paradoxically confer survival benefit in MO□ACLD. This aligns with the obesity paradox described in advanced cirrhosis [28], implying that, when adipose–muscle energy transfer remains functional, maintaining energy reserves could improve tolerance to nutritional stress and extend survival. This provides valuable insights into nutritional management and therapeutic strategies for advanced MASLD.

Our survival analysis (**Figure 6**) revealed a critical heterogeneity hidden within the broader categories of MP. This "meta-paradox" was clearly demonstrated when we compared the outcomes of three distinct patient groups within our ACLD cohort (excluding HCC). As detailed in **Supplementary Table 7**, these groups—non-burning-out MO, MP alone, and burning-out MO—exhibited a clear step-wise deterioration in baseline ALBI score. This hierarchy in liver function translated directly into their survival. A clear trend was observed in the 3-year survival rates: 91% for the non-burning-out MO group (n=17), 79% for the MP alone group (n=84), and 53% for the burning-out MO group (n=26), though these differences should be interpreted with caution given the small sample sizes.

Importantly, many patients classified as burning-out MO likely already exhibit early signs of liver failure. Longitudinal analysis suggests that coexistence of the burnout and liver-failure phenotypes represents a simultaneous collapse of energy supply and hepatic synthetic capacity. This composite phenotype characterizes end-stage MASLD and explains the rapid decline in survival, exceeding that observed in non-burning-out MO or MP alone.

This tiered outcome provides two critical insights. First, the superior survival in the non-burning-out MO group compared with the MP-alone group clearly demonstrates the "obesity paradox." Second, and more importantly, the primary determinant of prognosis appears to be liver functional reserve rather than obesity per se.

When hepatic energy supply pathways fail, obesity loses its beneficial effect, resulting in the poorest prognosis. Indeed, our cross-sectional analysis clearly demonstrated that reduced liver fat content and metabolic collapse (burning-out) closely correlate with rapid deterioration in hepatic functional reserve.

This finding is extremely important as it suggests that the pathophysiology of the obesity-MASLD-sarcopenia cascade is fundamentally different from muscle loss and cachexia caused by other chronic wasting diseases such as renal failure, heart failure, and cancer. Our results indicate that in MASLD, the breakdown of hepatic metabolic function may be a central and rate-limiting event driving terminal catabolism, thereby uniquely defining the “sarcopenia” observed in CLD.

### 3. A Self-Perpetuating Metabolic Loop

In our obese patient population with early-stage fibrosis (F0–2), improvement in hepatic steatosis (defined as a decrease in PDFF ≥ 1%) was strongly associated with reductions in both VAT and muscle mass. These findings align with previous reports indicating that simple weight loss, regardless of method, often results in concurrent muscle loss [29, 30]. This underscores a critical metabolic trade-off: fat reduction remains a key therapeutic target in MASLD, yet frequently occurs at the expense of skeletal muscle, particularly without protective interventions. Crucially, our study provides, for the first time, direct statistical evidence validating this trade-off in early-stage MASLD. The comprehensive response screening analysis of our F0-2 cohort identified a significant, FDR-adjusted correlation between the annual change in mVAT and muscle mass (FDR-adjusted P = 0.048).

This muscle loss can be viewed as an adaptive response to transient starvation induced by caloric restriction. Under negative energy balance, the body mobilizes adipose tissue while simultaneously initiating muscle catabolism, reducing resting energy expenditure and providing amino acids for gluconeogenesis [31, 32]. However, this adaptation becomes maladaptive in MASLD, ultimately compromising metabolic health. Crucially, resultant MP is not merely collateral damage—it actively fuels a self-perpetuating metabolic loop by impairing insulin sensitivity, accelerating metabolic dysfunction, and promoting hepatic re-steatosis. This loop is particularly detrimental when fat loss occurs without muscle preservation, widening the fat–muscle gap and exacerbating metabolic imbalance.

The clinical significance extends beyond disease progression, critically impacting post-transplant outcomes. MASLD is currently a leading indication for liver transplantation (LT), with registry data indicating hepatic re-steatosis in approximately 60% of recipients within one year and over 80% within five years post-transplant [33, 34]. This study identifies VAT as the primary upstream trigger of the MASLD cascade, and impaired liver reserve with reduced PDFF ("burning-out") as the hallmark of advanced MASLD. Although LT dramatically restores hepatic function, it inadvertently resets conditions favoring re-initiation of this metabolic loop—akin to reopening a previously closed floodgate, permitting rapid energy influx into the liver and potentially reactivating the obesity–MASLD cascade.

### 4. Stage-specific treatment strategies: Breaking out of the metabolic loop

Our findings reveal that escaping the obesity–MASLD–myopenia cascade demands fundamentally distinct therapeutic strategies at each disease stage—a therapeutic metamorphosis mirroring the metabolic transformation itself.

In the early stages (F0–2), VAT represents the primary metabolic threat. Currently, GLP-1 receptor agonists (GLP-1RAs) have garnered significant attention due to their potent fat-reducing effects. However, GLP-1RAs frequently induce concomitant skeletal muscle loss [35], raising concerns regarding their long-term metabolic safety. In our cohort, although not statistically significant, patients on antidiabetic agents—including GLP-1RAs (n=7)—exhibited a trend toward greater annual muscle loss than non-users (–3.1% vs. –2.4%, *P*=0.45). This finding suggests that selective fat reduction must be strategically balanced with muscle preservation, as unintended muscle loss may compromise metabolic resilience and predispose patients to rebound weight gain. Therefore, treatment in this stage should prioritize high-quality weight loss, defined as targeted VAT reduction with minimal muscle loss. This goal may be best achieved through a combination of resistance training and high-protein nutritional support.

A critical limitation of current clinical guidelines is their reliance on total body weight as the primary therapeutic target, without adequately addressing the essential aspect of muscle preservation. Simply losing weight without actively preserving muscle mass may paradoxically accelerate progression into the metabolic loop. If such qualitative weight loss can be achieved, the current MASLD guideline targets—based solely on total body weight reduction—may be relaxed.

Conversely, as disease advances into cirrhosis, the therapeutic paradigm shifts drastically—from energy reduction strategies to aggressive energy repletion—as the clinical challenge transitions from excess energy stores to severe energy deficits. Two recent randomized controlled nutrition trials in cirrhotic patients [36, 37] demonstrated that branched-chain amino acid supplementation alone failed to improve muscle mass and strength, whereas adding approximately 1,300 kcal/day of extra energy significantly increased lean body mass, handgrip strength, and notably, immune function. Weight loss at this critical juncture is tantamount to being counterproductive.

The challenge of nutritional therapy in advanced MASLD lies in the gradual nature of this Metabolic Metamorphosis. In this longitudinal analysis, worsening ALBI scores in patients at stages F3-4 were significantly associated with decreases in muscle mass, body fat mass, and liver fat mass ("burning-out"), while increases in body fat mass were associated with elevated PDFF and ALT levels ("burn-up"). This indicates a complex, mixed state of anabolism and catabolism, reflecting the coexistence of these contrasting metabolic states. Therefore, it is essential to establish individualized nutritional therapy goals for each patient.

Post-LT introduces the ultimate paradox: the newly functional liver can reset conditions for cascade recurrence, yet muscle mass recovery is not expected [38]. Our F3-4 data linking ALBI score to muscle mass reinforce that myopenia is an irreversible change requiring active intervention. The obesity-MASLD-myopenia cascade is thus a self-perpetuating loop that amplifies muscle loss, a process continuing even post-transplantation. In essence, muscle loss is a one-way ticket, underscoring why immediate, multidisciplinary intervention focused on aggressive muscle preservation and metabolic management is essential to break this vicious cycle.

## Limitations

### 1. Generalizability and lifestyle confounding

All participants were East-Asian patients from a single tertiary center. Data on physical activity, diet, and other lifestyle factors were not collected, limiting extrapolation to other populations and preventing adjustment for lifestyle-related confounding.

### 2. Selection bias toward compensated disease

Inclusion was limited to patients able to undergo MRE, excluding those with advanced decompensation or severe comorbidities. This may limit representation of the full disease spectrum, though it reduced confounding and enabled clearer stage-specific characterization. MRE timing and frequency were determined by clinical judgment rather than standardized protocols, resulting in variable follow-up intervals. Such variability may have influenced muscle-loss estimates, especially in early-stage fibrosis (F0–2), where shorter follow-up was associated with higher loss rates.

**Supplementary Table 8** suggests that the variability in follow-up intervals may reflect differences in treatment goals for MASLD and surveillance needs for viral hepatitis, such as HCC risk. The higher rate of muscle loss in the short-term group could represent the initial, aggressive phase of lifestyle modification, where the primary goal of improving liver parameters may be achieved at the expense of muscle mass. In contrast, the long-term group might be enriched with a selected cohort requiring ongoing management due to treatment resistance or higher intrinsic risk (e.g., post-HCV cirrhosis). This heterogeneity, while a limitation, uniquely mirrors real-world clinical practice, where patient management is tailored and not standardized. This highlights complexities that might not be captured in prospective trials with rigid, standardized imaging intervals.

### 3. Lack of functional muscle assessment

Our MRI-based methodology focused on muscle *mass*, a key metabolic parameter, rather than function. Thus, our findings concern myopenia and are not directly comparable to studies using full sarcopenia criteria. Future work should link these quantitative changes in mass to functional impairment.

### 4. Causality and follow-up

Although a longitudinal cohort allowed modeling of parameter changes, the retrospective design and variable follow-up durations limit causal inference. Observed associations should be viewed as a strong hypothesis for the obesity–MASLD–myopenia cascade, requiring confirmation in prospective studies with predefined intervals.

### 5. Drug therapy effects

Short follow-up may have underestimated the impacts of GLP-1 receptor agonists and SGLT2 inhibitors on muscle mass.

Despite these limitations, the consistency of our findings across cross-sectional and longitudinal analyses, together with their alignment with established pathophysiological mechanisms, likely represents a valid framework for understanding the obesity–MASLD–myopenia cascade.

## Conclusions

This study proposes and describes the obesity–MASLD–MP cascade, a conceptual framework redefining MASLD as a dynamic metabolic metamorphosis and loop. Persistent muscle loss within this cascade may be a critical determinant of poor outcomes, independent of obesity, suggesting that therapeutic priorities should evolve with disease stage. The self-perpetuating loop may be the primary driver underlying this metabolic cascade.

In late-stage MASLD, the “burning-out” phenotype with muscle loss appears shaped by portal-hypertensive hemodynamic rerouting and hepatic dysfunction. Functionally, the liver becomes an “isolated island” in whole-body energy metabolism—cut off from VAT-derived substrates and signals—failing to participate in the VAT-centered obesity paradox that may still benefit extrahepatic organs. Early intervention should be considered important to prevent entry into this potentially irreversible metabolic spiral.

## Data Availability

All data produced in the present study are available from the corresponding author upon reasonable request.

## Acknowledgments

None.

## Author contributions

TI, KO: study concept and design; TI, KO: patient recruitment and characterisation; TI, KO: data acquisition; TI, KO: data analysis; TI, KO: article drafting. All authors provided input and critical revision and approved the final version.

