## Supplemental files for "The Obesity–MASLD–Myopenia Cascade: A Dynamic Loop within Metabolic Metamorphosis in Liver Disease"

**Supplementary Material**

### 1. Methods

- Methods (1)–(5)

### 2. Supplementary Information

- Tables 1–8
- Figures 1–2

**1.1 Disease Classification, Staging, and Imaging Measurements**

Steatotic liver disease (SLD) was diagnosed based on imaging findings (hepatic and renal contrast on abdominal ultrasonography; liver/spleen ratio < 0.9 on abdominal computed tomography (CT); proton density fat fraction measured by magnetic resonance imaging (MRI), > 5.2%[1]. Additionally, alcohol-related liver disease was diagnosed in accordance with the diagnostic criteria of the Japanese Society for Biomedical Research on Alcohol [2], and metabolic dysfunction-associated steatotic liver disease (MASLD) was diagnosed according to the new diagnostic criteria[3]. Hepatocellular carcinoma (HCC) was diagnosed according to the guidelines as tumors marked in the arterial phase on contrast-enhanced CT and showing washout in the portal or delayed phase[4,5]. In the case of gadoxetate disodium (EOB) contrast-enhanced MRI, tumors were also defined as those that stained in the early phase and showed washout in the portal venous phase.

**1.2** **Diagnostic criteria for advanced CLD (ACLD), and procedures for diagnosing the severity of hepatic reserve dysfunction:** The diagnosis of ACLD in this study was based on LS ≥ 3kPa on magnetic resonance elastography (MRE) in accordance with the practice guidance by the American Association for the Study of Liver Diseases (AASLD)[6]. Liver reserve function impairment was evaluated using the albumin-bilirubin (ALBI) score, and disease severity was categorized based on ALBI grade (grade 1 to 3) and the Model for End-Stage Liver Disease–sodium (MELD-Na) score[7,8].

All patients underwent liver MRI after fasting for more than 12 hours, using a 1.5-T whole-body scanner (SIGNA Voyager XT 1.5T; GE Healthcare, Tokyo, Japan). The imaging protocol included axial and coronal T1- and T2-weighted images with fat suppression, diffusion-weighted imaging, and magnetic resonance elastography (MRE) using 60 Hz acoustic vibration delivered via a pneumatic driver positioned over the right thorax. Liver stiffness (LS) was measured by MRE. A 19 cm passive pneumatic driver connected to an acoustic waveform generator transmitted 60 Hz vibrations to the right thorax at the xiphoid level. Wave propagation was captured using a gradient-echo sequence, and the region of interest (ROI) was carefully placed in the right hepatic lobe, avoiding large vessels, bile ducts, gallbladder, tumors, and imaging artifacts. LS values (in kPa) were assessed by experienced radiologists. Intrahepatic fat content was quantified using the IDEAL IQ sequence to calculate proton density fat fraction (PDFF), expressed as a percentage. All imaging-derived quantitative measurements were conducted by a single trained radiologist with expertise in clinical hepatology trials. Both LS and PDFF values were used in subsequent analyses as quantitative indicators of liver fibrosis and steatosis, respectively.

Fibrosis staging was determined using etiology-specific MRE-LS cutoffs.
For MASLD, fibrosis stages were defined according to Hsu et al. [9] as follows:

- Stage 0: LS < 2.61 kPa
- Stage 1: 2.61–2.96 kPa
- Stage 2: 2.97–3.61 kPa
- Stage 3: 3.62–4.68 kPa
- Stage 4: ≥ 4.69 kPa

For non-MASLD etiologies, fibrosis staging followed Morisaka et al. [10]:

- Stage 0: LS < 2.32 kPa
- Stage 1: 2.32–2.60 kPa
- Stage 2: 2.61–3.01 kPa
- Stage 3: 3.02–4.22 kPa
- Stage 4: ≥ 4.23 kPa

Among patients with stage 4 fibrosis, cirrhosis was further classified as compensated (cLC) or decompensated (dLC), based on the presence of major clinical events such as ascites, hepatic encephalopathy, or gastrointestinal bleeding from varices [11].

**1.3 MRI-Based Quantification and Diagnostic Criteria**

S1. **Diagnostic Criteria** for Obesity, Myopenia, and Myopenic Obesity

**・Obesity.** Obesity was defined as a body mass index (BMI) ≥ 25 kg/m² according to the World Health Organization (WHO) Asia–Pacific classification [12].

**・Myopenia (MP).** Skeletal muscle mass was quantified on 1.5-T MRI as previously described [13,14]. Paraspinal muscle area (PSMA) was traced on T2-weighted single-shot fast spin-echo images at the level of the superior mesenteric artery (SMA). Bilateral erector spinae, multifidus, quadratus lumborum, and spinalis muscles were summed and height-adjusted to yield the paraspinal muscle index (PSMI, cm²/m²). Sex-specific cut-offs (male < 12.62 cm²/m², female < 9.77 cm²/m²) identified MP. Repeatability was not re-evaluated in the present study, as our previous work had already confirmed excellent intra- and inter-observer reproducibility for this single-slice protocol [14].

**・Myopenic obesity (MO).** MO was diagnosed when both obesity and MP were resent.

S2. MRI-Based Quantification of Body Composition

MRI was chosen for its ability to assess skeletal muscle mass, intra-hepatic fat (PDFF), and liver stiffness (LS) in a single session, offering superior accuracy over CT or ultrasonography.

**Single-slice protocol.** Adipose tissue and muscle were measured on an axial T1-weighted slice at the SMA origin (**Supplementary Fig. 1**). This landmark is highly reproducible, allowing consistent positioning across serial scans.

**Rationale and robustness.** The limited z-coverage of MR elastography often precludes whole-abdomen segmentation. Our standardized single-slice method minimizes positional variability and yields robust, reproducible estimates of longitudinal change. Annualized percentage change (Δ % / year) was calculated for:

- visceral adipose tissue (VAT)
- subcutaneous adipose tissue (SAT)
- skeletal muscle mass (PSMA)

**Limitations.** VAT could not be quantified in the presence of overt ascites; therefore, adipose-tissue analyses were restricted to compensated patients.

### S3. Justification for Muscle-Mass–Based MP Definition

### Current AASLD guidance endorses imaging-based assessment of sarcopenia. Muscle strength tests (e.g., hand-grip) were excluded because they are influenced by non-musculoskeletal factors and do not directly reflect the energy-storage function of muscle, which was central to our metabolic trade-off hypothesis involving adipose depots and liver fat.

**1.4 Outcome and Follow-up**

Patients with advanced chronic liver disease (ACLD: LS ≥ 3 kPa) were followed from the date of baseline MRI to the date of liver-related death, liver transplantation, last clinic visit, or confirmation of survival. For patients who underwent liver transplantation, the date of transplant surgery was considered the censoring point. The primary outcome of the study was liver-related mortality, defined as death attributable to hepatic decompensation, variceal bleeding, spontaneous bacterial peritonitis, or hepatocellular carcinoma. Patient outcomes were initially reviewed through electronic medical records. For patients who were referred to other institutions, outcome data were obtained via interinstitutional communication. In cases where outcomes remained uncertain, direct contact with the patient or their family was attempted by phone. The median follow-up period was 34.0 months (interquartile range: 3–73 months). Patients with incomplete follow-up information were censored at the date of last confirmed contact.

**1.5 Statistical Analysis**

All statistical analyses were performed using JMP version 18.2.0 (SAS Institute Japan). Between-group differences were assessed using the chi-square test for categorical variables and the Mann–Whitney–Wilcoxon test for continuous variables. For comparisons among three or more groups, the Kruskal–Wallis test was used, followed by post hoc Steel–Dwass tests to identify significant pairwise differences.

Survival analysis in patients with ACLD was conducted using the Kaplan–Meier method, with differences between groups assessed by the log-rank test. A Cox proportional hazards model was employed to identify independent prognostic factors. For multivariate analysis, variables were initially selected using stepwise forward and backward methods, and a minimal set of clinically relevant factors was retained to construct the final model. Given the exploratory nature of univariate and subgroup comparisons, no correction for multiple testing was applied. In all analyses, a p-value of < 0.05 was considered statistically significant.

**References**

1. Imajo K, Kessoku T, Honda Y, et al. Magnetic resonance imaging more accurately

classifies steatosis and fibrosis in patients with nonalcoholic fatty liver disease than

transient elastography. Gastroenterology 2016; 150: 626–37.

2. Takada A, Matsuda Y, Takase S, et al. A national surveillance study on alcoholic

liver disease in Japan (1986–1991). Nihon Shokakibyo Gakkai Zasshi 1994; 91:

887–98.

3. Rinella ME, Lazarus JV, Ratziu V, et al. A multisociety Delphi consensus statement on new fatty liver disease nomenclature. Hepatology 2023; 78: 1966-1986.

4. Marrero JA, Kulik LM, Sirlin CB, et al. Diagnosis, staging, and management of

hepatocellular carcinoma: 2018 practice guidance by the American Association for

the Study of Liver Diseases. Hepatology 2018; 68: 723–50.

5. Kokudo N, Takemura N, Hasegawa K, et al. Clinical practice guidelines for

hepatocellular carcinoma: the Japan Society of Hepatology 2017 (4th JSH- HCC

guidelines) 2019 update. Hepatol Res 2019; 49: 1109–13.

6. J Kaplan DE, Bosch J, Ripoll C, et al. AASLD practice guidance on risk

stratification and management of portal hypertension and varices in cirrhosis.

Hepatology 2023. doi: 10.1097/HEP.0000000000000647

7. Johnson PJ, Berhane S, Kagebayashi C, et al. Assessment of liver function in patients with hepatocellular carcinoma: a new evidence-based approach-the ALBI grade. J Clin Oncol 2015;33:550-8.

8. W Ray Kim, Scott W Biggins, Walter K Kremers, et al. Hyponatremia and mortality among patients on the liver-transplant waiting list. N Engl J Med 2008;359:1018-26.

9. Hsu C, Caussy C, Imajo K, et al. Magnetic Resonance vs Transient Elastography Analysis of Patients With Nonalcoholic Fatty Liver Disease: A Systematic Review and Pooled Analysis of Individual Participants. Clin Gastroenterol Hepatol 2019;17:630-637.e8. doi: 10.1016/j.cgh.2018.05.059.
10. Morisaka H, Motosugi U, Ichikawa S, et al. Magnetic resonance elastography is as accurate as liver biopsy for liver fibrosis staging. J Magn Reson Imaging 2018;47:1268-1275. doi: 10.1002/jmri.25868.
11. European Association for the Study of the Liver. EASL Clinical Practice Guidelines for the management of patients with decompensated cirrhosis. J Hepatol 2018;69:406-460.
12. The Asia-Pacific perspective: redefining obesity and its treatment.
    Sydney: Health Communications Australia; 2000. Available from: <https://iris.who.int/handle/10665/206936>
13. Praktiknjo M, Book M, Luetkens J, Pohlmann A, Meyer C, Thomas D, et al. Fat-free muscle mass in magnetic resonance imaging predicts acute-on-chronic liver failure and survival in decompensated cirrhosis. Hepatology 2018;67:1014-1026. doi: 10.1002/hep.
14. Nakamura A, Yoshimura T, Sato T, et al. Diagnosis and Pathogenesis of Sarcopenia in Chronic Liver Disease Using Liver Magnetic Resonance Imaging. Cureus 2022;14:e2467 doi: 10.7759/cureus.24676
15. Lai JC, Tandon P, Bernal W, et al. Malnutrition, Frailty, and Sarcopenia in Patients With Cirrhosis: 2021 Practice Guidance by the American Association for the Study of Liver Diseases. Hepatology 2021 ;74:1611-1644.

**Supplementary Table 1**

Comparison of PDFF, ALT, and ALBI scores across fibrosis stages in MASLD and non-MASLD groups. Data are presented as median (interquartile range).

| **factors** | **F stage** | **MASLD** | **Non-MASLD** | **P-value** |
| --- | --- | --- | --- | --- |
| **PDFF (%)** | **F0-1** | 11 (7-17) | 3 (2-6) | <0.001 |
|  | **F2-3** | 15 (9-20) | 4 (2-7) | <0.001 |
|  | **F4c** | 13 (7-21) | 4 (2-7) | <0.001 |
|  | **F4d** | 4 (2-7) | 3 (2-5) | 0.322 |
| **ALT (IU/L)** | **F0-1** | 39 (22-70) | 18 (13-29) | <0.001 |
|  | **F2-3** | 62 (42-94) | 28 (18-45) | <0.001 |
|  | **F4c** | 50 (35-104) | 35 (22-63) | 0.021 |
|  | **F4d** | 25 (15-30) | 28 (18-42) | 0.314 |
| **ALBI score** | **F0-1** | -3.05 (-3.26 to -2.85) | -3.12 (-3.31 to -2.92) | 0.089 |
|  | **F2-3** | -2.89 (-3.10 to -2.64) | -2.95 (-3.19 to -2.71) | 0.182 |
|  | **F4c** | -2.58 (-2.78 to -2.31) | -2.65 (-2.84 to -2.38) | 0.243 |
|  | **F4d** | -1.65 (-2.18 to -1.34) | -1.56 (-1.92 to -1.06) | 0.526 |

**PDFF**, proton density fat fraction; **ALT**, alanine aminotransferase; **ALBI score**, albumin-bilirubin score; **MASLD**, metabolic dysfunction-associated steatotic liver disease; **Non-MASLD**, chronic liver disease other than MASLD; **F stage**, fibrosis stage (F0–1, no significant fibrosis; F2–3, significant fibrosis without cirrhosis; F4c, compensated cirrhosis; F4d, decompensated cirrhosis).
Statistical significance was assessed using the Mann–Whitney U test.

**Supplementary Table 2**

Longitudinal Clinical Profile in Patients with BMI ≥ 25 kg/m²: MASLD vs. Non-MASLD Comparison

|  | **all** | **MASLD** | **Non MASLD** | P-value |
| --- | --- | --- | --- | --- |
| (Number) | **(169)** | **(84)** | **(85)** |  |
| Observation interval (months) | 39.1±19.7 | 35.6±20.2 | 42.6±18.8 | * |
| Age (y) | 62±13 | 59±15 | 64±10 | * |
| Sex (M/F) | 114/55 | 53/31 | 61/24 |  |
| BMI (kg/m^2^) | 28±9 | 29±3 | 27±3 | * |
| ***Etiology of Liver Disease*** |  |  |  |  |
| MASLD | 84 (50) | 84 (100) | 0 (0) |  |
| Hepatitis B | 23 (14) | 0 (0) | 23 (27) |  |
| Hepatitis C | 39 (23) | 0 (0) | 39 (46) |  |
| ALD | 14 (8) | 0 (0) | 14 (16) |  |
| Others | 9 (5) | 0 (0) | 9 (11) |  |
| ***Laboratory data*** |  |  |  |  |
| Baseline AST (U/L) | 40±28 | 45±31 | 35±24 | * |
| Baseline  ALT (U/L) | 51±55 | 62±50 | 39±57 | * |
| Final AST (U/L) | 33±18 | 36±19 | 32±18 |  |
| Final ALT (U/L) | 34±25 | 37±25 | 31±24 | * |
| **ALT change (ΔU/l/year)** | -8±22 | -12±23 | -4±20 | * |
| Baseline LS (kPa) | 3.62±2.12 | 3.38±1.99 | 3.86±2.23 | * |
| Final LS (kPa) | 3.37±1.56 | 3.10±1.47 | 3.63±1.61 | * |
| **LS change (Δ kPa/year)** | -0.19±1.07 | -0.22±0.90 | -0.15±1.21 |  |
| Baseline  ALBI score | -2.78±0.45 | -2.87±0.56 | -2.72±0.51 | * |
| Final ALBI score | -2.79±0.53 | -2.87±0.37 | -2.69±0.50 | * |
| **ALBI change (Δ /year)** | -0.03±0.39 | -0.01±0.29 | -0.06±0.47 |  |
| Baseline PDFF (%) | 11.8±8.2 | 15.5±8.4 | 8.2±6.2 | * |
| Final PDFF (%) | 11.1±8.2 | 13.8±8.3 | 7.3±7.3 | * |
| **PDFF change (Δ %/year)** | -0.2±2.5 | -0.6±3.2 | 0.2±1.4 | * |
| Baseline HbA1c (%) | 6.1±1.0 | 6.3±1.1 | 6.0±0.9 |  |
| Final HbA1c (%) | 6.2±0.9 | 6.3±0.9 | 6.1±0.8 |  |
| ***Body composition*** |  |  |  |  |
| SAT change (Δ %/year) | 0.3±5.1 | 0.4±6.1 | 0.1±3.9 |  |
| VAT change (Δ %/year) (n=164) | 0.6±4.6 | 0.7±5.6 | 0.6±3.4 |  |
| Muscle change (Δ %/year) | -2.5±4.4 | -3.2±5.0 | -1.8±3.5 |  |
| Baseline myopenia (n) | 26 | 7 | 19 | * |
| Final myopenia (n) | 38 | 11 | 27 | * |

Data are presented as mean ± SD or number (%).
BMI, body mass index; MASLD, metabolic dysfunction-associated steatotic liver disease; ALD, alcohol-associated liver disease; AST, aspartate aminotransferase; ALT, alanine aminotransferase; LS, liver stiffness; ALBI, albumin–bilirubin; PDFF, proton density fat fraction; HbA1c, hemoglobin A1c; SAT, subcutaneous adipose tissue; VAT, visceral adipose tissue.
*p <0.05 indicate statistically significant differences between MASLD and Non-MASLD groups.
Patients included had a BMI ≥ 25 kg/m².

**Supplementary Table 3**

Spearman’s correlation analyses examining relationships among annual changes in body composition (muscle and fat), liver fat content (PDFF), ALT, liver stiffness (LS), and final HbA1c values in patients with liver fibrosis stage **F0–2 (n=103)**.

Data presented as correlation coefficients (Spearman’s ρ) with corresponding P-values.
(1) Correlations between annual muscle volume change and other parameters.
(2) Correlations among annual changes in fat mass, PDFF, ALT, and LS.
(3) Correlations between final HbA1c values and annual changes in metabolic and liver-related parameters.

**(1)**

| **Variable Pair** | **Spearman’s ρ** | **P-value** |
| --- | --- | --- |
| Annual muscle change (%/y) vs. Annual fat change (%/y) | 0.126 | 0.204 |
| Annual muscle change (%/y) vs. Annual VAT (%/y) | 0.213 | 0.030 |
| Annual muscle change vs. Annual PDFF (%/y) | 0.051 | 0.608 |
| Annual muscle change vs. Annual ALT (U/l/y) | 0.028 | 0.779 |
| Annual muscle change vs. Annual LS (kPa/y) | 0.101 | 0.309 |

**(2)**

| **Variable Pair** | **Spearman’s ρ** | **P-value** |
| --- | --- | --- |
| Annual fat change vs. Annual PDFF | 0.505 | <0.001 |
| Annual fat change vs. Annual ALT | 0.423 | <0.001 |
| Annual fat change vs. Annual LS | 0.280 | 0.004 |
| Annual PDFF change vs. Annual ALT | 0.401 | <0.001 |
| Annual PDFF change vs. Annual LS | 0.144 | 0.146 |
| Annual ALT change vs. Annual LS | 0.432 | <0.001 |

**(3)**

| **Variable Pair** | **Spearman’s ρ** | **P-value** |
| --- | --- | --- |
| Last HbA1c (%) vs. Annual muscle change (%/y) | 0.001 | 0.993 |
| Last HbA1c (%) vs. Annual fat (%/y) | 0.245 | 0.017 |
| Last HbA1c (%) vs. Annual VAT (%/y) | 0.280 | 0.006 |
| Last HbA1c (%)  vs. Annual PDFF (%/y) | 0.073 | 0.485 |
| Last HbA1c (%) vs. Annual ALT (U/l/y) | -0.060 | 0.566 |
| Last HbA1c (%)   vs. Annual LS (kPa/y) | -0.004 | 0.965 |

ALT, alanine aminotransferase; PDFF, proton density fat fraction; VAT, visceral adipose tissue; LS, liver stiffness; HbA1c, hemoglobin A1c.
Statistical significance: P-values <0.05.

**Supplementary Table 4**

Spearman’s correlation analyses examining relationships **among** annual changes in body composition (muscle and fat), liver fat content (PDFF), ALT, liver stiffness (LS), and final HbA1c values **among** patients with liver fibrosis stage **F0–2 with MASLD (n=63)**.

Data presented as correlation coefficients (Spearman’s ρ) with corresponding P-values.
(1) Correlations between annual muscle volume change and other parameters.
(2) Correlations among annual changes in fat mass, PDFF, ALT, and LS.
(3) Correlations between final HbA1c values and annual changes in metabolic and liver-related parameters.

**(1)**

| **Variable Pair** | **Spearman’s ρ** | **P-value** |
| --- | --- | --- |
| Annual muscle change (%/y) vs. Annual fat change (%/y) | 0.099 | 0.442 |
| Annual muscle change (%/y) vs. Annual VAT (%/y) | 0.259 | 0.039 |
| Annual muscle change vs. Annual PDFF (%/y) | -0.033 | 0.800 |
| Annual muscle change vs. Annual ALT (U/l/y) | 0.039 | 0.767 |
| Annual muscle change vs. Annual LS (kPa/y) | 0.188 | 0.141 |

**(2)**

| **Variable Pair** | **Spearman’s ρ** | **P-value** |
| --- | --- | --- |
| Annual fat change vs. Annual PDFF | 0.316 | 0.012 |
| Annual VAT change vs. Annual PDFF | 0.505 | <0.001 |
| Annual fat change vs. Annual ALT | 0.480 | <0.001 |
| Annual VAT change vs. Annual ALT | 0.467 | <0.001 |
| Annual fat change vs. Annual LS | 0.288 | 0.022 |
| Annual VAT change vs. Annual LS | 0.247 | 0.051 |
| Annual PDFF change vs. Annual ALT | 0.427 | <0.001 |
| Annual PDFF change vs. Annual LS | 0.155 | 0.224 |
| Annual ALT change vs. Annual LS | 0.493 | <0.001 |

**(3)**

| **Variable Pair** | **Spearman’s ρ** | **P-value** |
| --- | --- | --- |
| Last HbA1c (%) vs. Annual muscle change (%/y) | 0.038 | 0.774 |
| Last HbA1c (%) vs. Annual fat (%/y) | 0.257 | 0.049 |
| Last HbA1c (%) vs. Annual VAT (%/y) | 0.291 | 0.026 |
| Last HbA1c (%)  vs. Annual PDFF (%/y) | 0.146 | 0.272 |
| Last HbA1c (%) vs. Annual ALT (U/l/y) | 0.123 | 0.354 |
| Last HbA1c (%)   vs. Annual LS (kPa/y) | 0.001 | 0.999 |

ALT, alanine aminotransferase; PDFF, proton density fat fraction; VAT, visceral adipose tissue; LS, liver stiffness; HbA1c, hemoglobin A1c.
Statistical significance: P-values <0.05.

**Supplementary Table 5**

Spearman’s correlation analysis of factors associated with annual changes in muscle volume, fat mass, liver fat (PDFF), ALT, liver stiffness and ALBI score in patients with liver fibrosis stage **F3-4 (n=66)**

**(1)**

| **Variable Pair** | **Spearman’s ρ** | **P-value** |
| --- | --- | --- |
| Annual ALBI score change vs. Annual muscle (%/y) | -0.269 | 0.029 |
| Annual ALBI score change  vs. Annual fat (%/y) | -0.308 | 0.017 |
| Annual ALBI score change vs. Annual PDFF (%/y) | -0.315 | 0.010 |
| Annual ALBI score change vs. Annual ALT (U/l/y) | -0.154 | 0.127 |
| Annual ALBI score change vs. Annual LS (kPa/y) | 0.092 | 0.465 |
| **(2)** |  |  |
| Annual muscle change (%/y) vs. Annual fat (%/y) | 0.215 | 0.083 |
| Annual muscle change vs. Annual PDFF (%/y) | 0.089 | 0.475 |
| Annual muscle change vs. Annual ALT (U/l/y) | -0.076 | 0.547 |
| Annual muscle change vs. Annual LS (kPa/y) | 0.022 | 0.863 |
| **(3)** |  |  |
| Annual fat change vs. Annual PDFF | 0.358 | 0.005 |
| Annual fat change vs. Annual ALT | 0.140 | 0.026 |
| Annual fat change vs. Annual LS | -0.118 | 0.371 |
| Annual PDFF change vs. Annual ALT | 0.316 | 0.010 |
| Annual PDFF change vs. Annual LS | 0.013 | 0.920 |
| Annual ALT change vs. Annual LS | 0.150 | 0.226 |

ALT, alanine aminotransferase; PDFF, proton density fat fraction; VAT, visceral adipose tissue; LS, liver stiffness; HbA1c, hemoglobin A1c.
Statistical significance: P-values <0.05.

**Supplementary Table 6**

Univariate analysis of factors associated with annual muscle volume loss stratified by liver fibrosis stage

| **Variable Pair** | **β** | **95%CI** | **P-value** |
| --- | --- | --- | --- |
| **1) Early stage (F0-2)** |  |  |  |
| Sex: Female (F0-2) | -0.204 | -0.160, -0.045 | 0.039 |
| BUN (F0-2) | 0.307 | 0.069, 0.514 | 0.011 |
| Cr (F0-2) | 0.212 | 0.383, 8.526 | 0.032 |
| Annual Δ VAT (%) (F0-2) | 0.206 | 0.011, 0.318 | 0.036 |
| Observation interval (months) (F0-2) | 0.333 | 0.030, 0.106 | 0.001 |
| **2) Advanced stage (F3-4)** |  |  |  |
| Age (F3-4) | -0.351 | -0.256, -0.051 | 0.004 |
| Annual Δ PDFF (%) (F3-4) | 0.510 | 0.722, 1.772 | < 0.001 |
| Baseline LS (F3-4) | -0.390 | -1.453, -0.376 | 0.001 |
| Baseline ALBI score (F3-4) | -0.262 | -4.772, -0.198 | 0.034 |
| Annual Δ ALBI score (F3-4) | -0.504 | -6.281, -2.514 | < 0.001 |

Univariate regression analysis examining factors associated with annual changes in muscle volume in patients stratified by liver fibrosis stage: early stage (F0–2) and advanced stage (F3–4). Data are presented as regression coefficients (β), 95% confidence intervals (CI), and corresponding P-values.
BUN, blood urea nitrogen; Cr, creatinine; VAT, visceral adipose tissue; PDFF, proton density fat fraction; LS, liver stiffness; ALBI, albumin–bilirubin.
Statistical significance was defined as P-value <0.05.

**Supplementary Table 7**

Comparison of clinical characteristics among MP alone, MO (PDFF ≥ 5%), and MO (PDFF < 5%) patients.

|  | **MP alone** | **MO**  **(PDFF** ≧ **5%)** | **MO**  **(PDFF < 5%)** |
| --- | --- | --- | --- |
| Patients number (n) | 84 | 17 | 19 |
| Age (yrs) | 69 ± 13 | 67 ± 10 | 68 ± 15 |
| Sex (Male) | 63% | 65% | 74% |
| BMI (kg/m^2^) | 21 ± 2 | 29 ± 6 * | 28 ± 3 * |
| PDFF (%) | 4.9 ± 5.4 | 10.8 ± 6.6 * | 2.2 ± 1.0 *† |
| ALBI score | -2.26 ± 0.70 | -2.40 ± 0.88 | -1.58 ± 0.66 *† |
| MELD Na | 10 ± 5 | 10 ± 7 | 14 ± 4 *† |

Data are presented as mean ± standard deviation or number (percentage).
MP, myopenia; MO, myopenic obesity; BMI, body mass index; PDFF, proton density fat fraction; ALBI, albumin–bilirubin; MELD Na, Model for End-Stage Liver Disease Sodium score.

* P< 0.05 vs. MP alone. † P< 0.05 vs. MO (PDFF ≥ 5%).

**Supplementary Table 8**

Comparison of clinical characteristics between short-term (< 39 months) and long-term (≥ 39 months) follow-up groups in obese patients with CLD (F0–2).

|  | **Short- Term Follow-up Group** | **Long-Term Follow-up Group** | **P-value** |
| --- | --- | --- | --- |
| ***Patient number*** | **56** | **47** |  |
| Observation interval (months) | 24.2±8.8 | 55.4±12.4 | ** |
| Age (y) | 59±15 | 59±10 |  |
| Sex (M/F) | 35/21 | 33/14 |  |
| BMI (kg/m^2^) | 28±3 | 27±3 |  |
| ***Etiology of Liver Disease*** |  |  |  |
| MASLD | 41 (73) | 22 (47) |  |
| Hepatitis B | 8 (14) | 6 (13) |  |
| Hepatitis C | 6 (11) | 12 (28) | ** |
| ALD | 1 (2) | 2 (4) |  |
| Others | 0 (0) | 4 (8) |  |
| ***Laboratory data*** |  |  |  |
| ALT change (ΔU/l/year) | -15±28 | -1±5 | ** |
| PDFF change (Δ %/year) | -0.51±3.49 | -0.04±1.50 |  |
| Hepatic fat reduction (ΔPDFF ≥ 1%/yr), n (%) | 21 (38) | 15 (32) |  |
| LS change (Δ kPa/year) | -0.04±0.29 | 0.08±0.15 | ** |
| ***Body composition*** |  |  |  |
| Muscle change (Δ %/year) | -3.1±4.7 | -1.3±2.1 | * |
| Baseline myopenia, n (%) | 5 (9) | 5 (11) |  |
| Final myopenia, n (%) | 11(20) | 10 (21) |  |
| VAT change (Δ %/year) | 0.3±6.3 | 1.5±1.9 | * |

Data are presented as mean ± SD or number (%). *: p < 0.05; **: p < 0.01. P-values were calculated using the Mann–Whitney U test for continuous variables and the χ² or Fisher’s exact test for categorical variables. Patients included had a BMI ≥ 25 kg/m².

BMI, body mass index; MASLD, metabolic dysfunction-associated steatotic liver disease; ALD, alcohol-associated liver disease; AST, aspartate aminotransferase; ALT, alanine aminotransferase; LS, liver stiffness; ALBI, albumin–bilirubin; PDFF, proton density fat fraction; HbA1c, hemoglobin A1c; SAT, subcutaneous adipose tissue; VAT, visceral adipose tissue.

**Supplementary Figures.**

**Supplementary Fig 1. Measurement of body composition changes using MRI.**

Representative MRI scans obtained at the level of the superior mesenteric artery (SMA) from the same patient at two time points (2020 and 2025). Outlined regions are mesenteric visceral adipose tissue (VAT; yellow dotted line), total adipose tissue (black dotted line), and the bilateral paraspinal muscle group (red solid line). Arrows highlight muscle (red), VAT (yellow), and subcutaneous adipose tissue (SAT; black).

**The consistent use of the SMA as an anatomical landmark ensured reproducible slice positioning across examinations, providing robust measurement accuracy and reliability.** This five-year comparison also demonstrates that, aside from muscle and adipose tissue, no major changes occurred in other abdominal organs, indicating that the measured differences reflect true changes in muscle mass, VAT, and SAT.

**Annualized percentage changes (Δ%) in visceral adipose tissue, total adipose tissue, and** bilateral paraspinal muscles **area were calculated using the following formula:**

**Annualized Δ% = [(Area at final scan − Area at baseline) / Area at baseline] ÷ follow-up duration (months) × 12 × 100.**

**Supplementary Figure 2.**

**Correlations between ALBI score, liver stiffness (LS), and liver fat content (PDFF) in patients with fibrosis stage F3–4 (without HCC)**

Scatterplots illustrating Spearman’s correlations between the ALBI score and (A) liver stiffness (LS, kPa) and (B) proton density fat fraction (PDFF, %), stratified by disease etiology (MASLD, orange circles; Non-MASLD, blue circles). Linear regression lines are shown for visual reference.

ALBI score correlated positively with LS in both MASLD and non-MASLD groups. However, the negative correlation between ALBI and PDFF differed by etiology: the non-MASLD group maintained consistently low PDFF values, whereas the MASLD group exhibited a marked decline in PDFF with decreasing liver functional reserve. Both groups ultimately converged toward a common metabolic endpoint characterized by hepatic fat loss.

Correlation coefficients (r) and associated P-values are indicated.

**Supplementary Figure 1.**
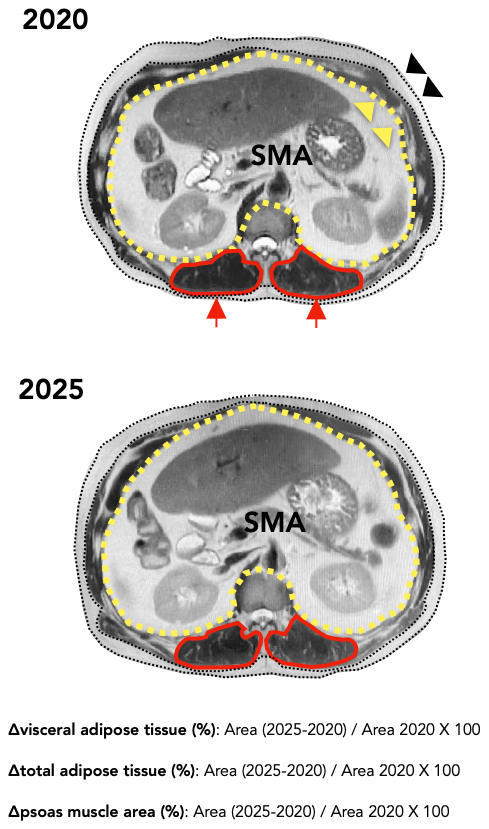

**Supplementary Figure 2.**
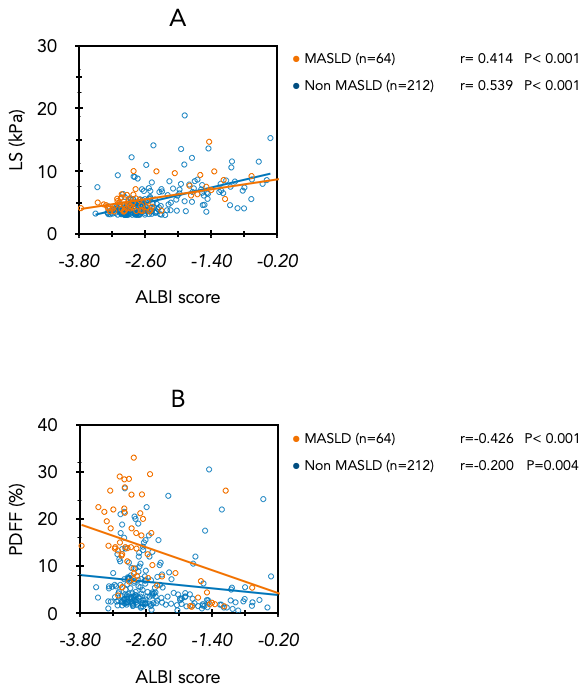
